# Repeated net-tDCS of the hypothalamus appetite-control network reduces inhibitory control and sweet food intake in persons with overweight or obesity

**DOI:** 10.1101/2024.12.11.24318873

**Authors:** Theresa Ester-Nacke, Ralf Veit, Julia Thomanek, Magdalena Book, Lukas Tamble, Marie Beermann, Dorina Löffler, Ricardo Salvador, Giulio Ruffini, Martin Heni, Andreas L. Birkenfeld, Christian Plewnia, Hubert Preissl, Stephanie Kullmann

**Author notes:** Corresponding author. Institute for Diabetes Research and Metabolic Diseases (IDM) of the Helmholtz Center Munich at the University of Tübingen. Otfried Müller Str. 47, 72076 Tübingen. E-mail address (T. Ester-Nacke).

## Abstract

**Background:** Reduced inhibitory control is associated with obesity and neuroimaging studies indicate that diminished prefrontal cortex activity influence eating behavior and metabolism. The hypothalamus regulates energy homeostasis and is functionally connected to cortical and subcortical regions especially the frontal areas.

**Objectives:** We tested network-targeted transcranial direct current stimulation (net-tDCS) to influence the excitability of brain regions involved in appetite control.

**Methods:** In a randomized, double-blind parallel group design, 44 adults with overweight or obesity (BMI 30.6 kg/m^2^, 52.3 % female) received active (anodal or cathodal) or sham 12-channel net-tDCS on the hypothalamus appetite-control network for 25 minutes on three consecutive days while performing a Stop-Signal-Task to measure response inhibition. Before and after stimulation, state questionnaires assessed changes in desire to eat and food craving. Directly after stimulation, participants received a breakfast buffet to evaluate *ad-libitum* food intake. An oral glucose tolerance test was conducted at follow-up. Resting-state functional MRI was obtained at baseline and follow-up.

**Results:** The Stop-Signal Reaction Time (SSRT) was shorter in both active groups versus sham, indicating improved response inhibition. Additionally, a stronger increase in hypothalamic functional connectivity was associated with shorter SSRT. Caloric intake of sweet food was lower in the anodal group versus sham, but no main effects between groups were observed on total and macronutrient intake, food craving ratings and desire to eat. At follow-up, no differences were observed between groups on peripheral metabolism.

**Conclusion:** Our study suggests that modulating hypothalamic functional network connectivity patterns via net-tDCS may improve food choice and inhibitory control.

**Graphical Abstract:** 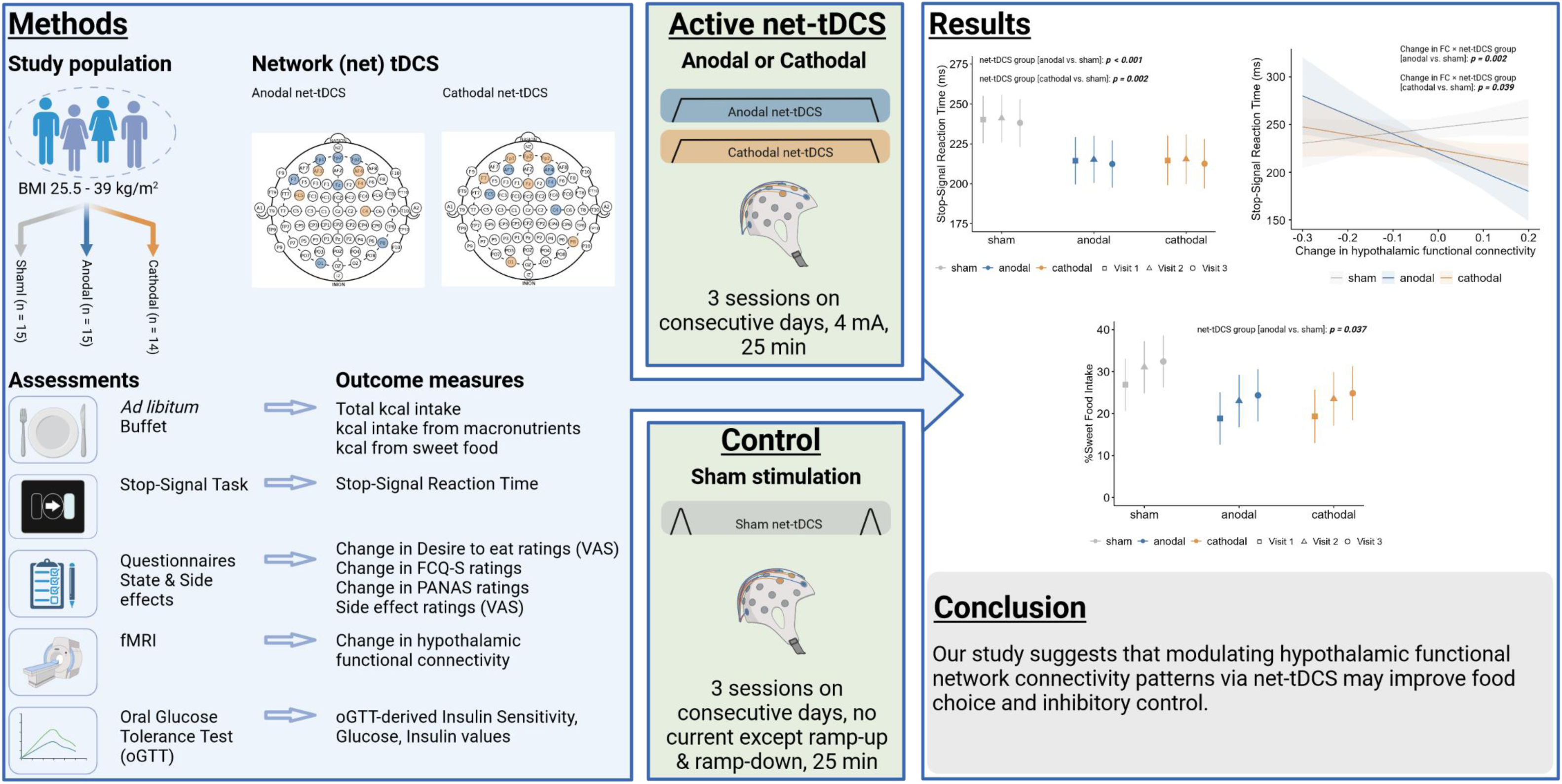

**Highlights:**

- Active net-tDCS groups showed better inhibitory control compared to the sham group.
- Stronger increase in hypothalamic functional connectivity associated with better inhibitory control after active net-tDCS.
- No differences were found between the active net-tDCS and sham groups for total kilocaloric intake.
- Anodal net-tDCS showed lower sweet food intake compared to the sham group.

## 1. Introduction

The prevalence of obesity nearly tripled between 1975 and 2016, representing one of the leading causes of morbidity and mortality [1]. Obesity is highly linked with an increased risk for numerous health conditions, notably cardiovascular diseases, insulin resistance and the development of type-2 diabetes (T2DM) [2]. Its etiology is complex and includes genetic, environmental, social and behavioral factors and the interactions among those [3] with a suspected significant contribution of the brain [4; 5].

In the long run, various obesity prevention and treatment approaches have demonstrated only limited success [2]. Among current interventions, bariatric surgery and pharmacological therapies represent the most effective approaches for significant weight loss and weight maintenance. Moreover, recent pharmacological therapies, particularly glucagon-like peptide 1 (GLP1) receptor agonists and glucose-dependent insulinotropic polypeptide (GIP)/GLP1 receptor co-agonists have shown promising results by targeting brain circuits involved in appetite control [6]. However, these interventions are not universally applicable and do not serve a preventive role, highlighting the need for new prevention approaches reducing obesity rates and maintain long-term weight management.

Thereby, the brain plays a pivotal role in regulating eating behavior wherein its altered response to peripheral hormones and external food cues increases the vulnerability to obesity [5; 7]. Central to this regulation is the hypothalamus, a key regulator of energy homeostasis, which is functionally connected to brain areas important for reward processing, food perception and cognitive control such as the prefrontal cortex (PFC), hippocampus and the striatum [8].

Neuroimaging studies in persons with overweight or obesity have identified modifications in these areas on a local and network level [for review [9–11]]. Specifically, the hypothalamus exhibits attenuated functional connectivity (FC) to brain regions crucial for cognitive control over food intake and increased FC to areas involved in reward [12; 13]. Moreover, persons with obesity show elevated food cue reactivity in brain regions important for reward and gustatory processing [14].

Inhibitory control refers to the ability to suppress a prepotent response (e.g. [15; 16]). Accordingly, impairments in dorsolateral PFC (dlPFC) activity are associated with reduced inhibitory control and higher impulsivity, which plays a role in the development and maintenance of obesity [17–19]. For instance, individuals with overweight or obesity showed lower FC between the hypothalamus and the dlPFC [13] and exhibit poorer performance in an inhibitory control task [20–22]. On the other hand, evidence from a functional magnetic resonance imaging (fMRI) study showed that increasing FC of the dlPFC enhanced control mechanisms related to food intake [23] and is related to better food choices [24]. Taken together, findings suggest that the brain activation is a predictor for dietary success [25] and understanding these connections offers profound insights into the neurobiological underpinnings of dietary habits and obesity.

Changing excitability of these brain areas may result in the modulation of food intake behavior and cognitive functions. Transcranial direct current stimulation (tDCS) serves as a non-invasive tool for neurostimulation to alter spontaneous firing rate and excitability of cortical neurons in the brain [26] to increase (anodal) or decrease (cathodal) resting membrane potential in neurons [27].

Several studies have shown that tDCS enhanced inhibitory control [28–30] reduced food craving and binge-eating episodes [31–37], especially for sweet foods [33; 35; 36; 38], lowered food intake [31; 39] and improved glucose metabolism [40–42]. Despite encouraging results, not all studies showed an effect of tDCS on food-related outcomes [for review see [43]], possibly due to inconsistency of stimulation parameters, large variation in experimental settings [44; 45] and inter-subject variations [46]. Moreover, previous efforts have mainly focused on stimulating isolated brain region, usually the dlPFC [43], without taking the interplay among various brain regions into account.

The hypothalamic FC network could be an ideal target for influencing eating behavior, reward and inhibitory control due to its functional connections to striatal and prefrontal regions. Using multiple small-sized electrodes, it is possible to target an entire brain network. This is referred to as network (net)-tDCS [47; 48]. Our recent pilot study [49] showed promising results with enhanced inhibitory control after anodal net-tDCS stimulation of this hypothalamus appetite-control network. This network covered areas of the dlPFC, ventromedial PFC, inferior frontal gyrus (IFG) and limbic regions that are critical for reward processes and decision making.

The current study examined net-tDCS targeting the hypothalamus appetite-control network to modify inhibitory control, food intake, food craving, and desire to eat in individuals with overweight or obesity and at high risk to develop T2DM. Given limited research on cathodal stimulation in this context, both anodal and cathodal forms were included. We hypothesized that the anodal net-tDCS group will show: (i) higher inhibitory control, as measured by the Stop-Signal Task (SST); (ii) lower caloric intake, food craving and desire to eat and; (iii) higher peripheral insulin sensitivity derived from an oral glucose tolerance test (oGTT) compared to those receiving sham or cathodal stimulation. For this purpose, we conducted a randomized, double-blind, placebo-controlled trial in parallel group design with resting-state fMRI as well as nutritional and psychometric evaluations before and after the 3-day net-tDCS stimulation and an oGTT at follow-up.

## 2. Materials and Methods

### 2.1 Study population

Men and women with overweight or obesity were recruited via email at the University Clinic Tübingen in Germany. Volunteers were eligible to participate if they met the following inclusion criteria: age between 20 to 65 years; body mass index (BMI) between 25.5 and 39 kg/m^2^ with a waist circumference of ≥ 80 cm in women and ≥ 94 cm in men; stable body weight (weight gain or loss ≤ 5 kg within 3 months). The exclusion criteria were: manifest T2DM; insufficient knowledge of the German language; pregnancy or lactation; history of severe mental or somatic disorders including neurological disease; drug or alcohol abuse; hemoglobin values ≤ 12g/dl for women and ≤ 14g/dl for men; magnetic resonance imaging (MRI) contra-indications; participation in a lifestyle intervention or pharmaceutical study. Written informed consent was obtained from all participants. The ethics committee of the University Hospital Tübingen approved the study protocol (project number: 243/2019BO1) and it was registered on ClinicalTrials-gov (registration number: NCT04420650). The trial was carried out from May 26, 2020, until September 28, 2023, according to the guidelines of the Declaration of Helsinki.

### 2.2 Experimental design and procedure

The study was conducted as a double-blind, parallel-group design, sham-controlled trial comparing active (anodal or cathodal) net-tDCS versus sham. The experimental design is depicted in **Fig. 1** (reporting checklist for tDCS studies is shown in **Suppl. Table 1**). In short, participants attended a total of five visits and were screened for eligibility criteria via phone prior to the baseline visit. At baseline, participants were interviewed for pre-existing psychiatric and neurological conditions during a detailed medical history assessment, followed by determination of body weight, body size, waist circumference and hip circumference. Participants underwent a resting-state fMRI and completed a series of psychometric questionnaires (see **Suppl. Methods** for questionnaire information). Following, three consecutive visits were conducted 7 to 76 days after the baseline visit. Participants were assigned using a block randomization method into either the anodal net-tDCS, cathodal net-tDCS or sham group. At the visits, participants, who were instructed to fast for 4-6 hours beforehand, arrived between 11:00 am and 3:00 pm and completed a battery of state questionnaires, namely Food Craving Questionnaire – State (FCQ-S), Desire to eat Visual Analogue Scale (VAS) and positive and negative affect schedule (PANAS) (pre net-tDCS, see **Suppl. Methods** for questionnaire information). This was followed by 25 minutes of either anodal, cathodal, or sham net-tDCS. During stimulation, participants completed a SST for 20 minutes. Subsequently, they repeated the state questionnaire battery (post net-tDCS) and filled out a questionnaire evaluating any side effects that may have occurred during stimulation. After the net-tDCS treatment, participants were left alone in an examination room where an *ad libitum* buffet was provided for 30 minutes. The follow-up took place one day after the last visit and included the same procedures as the baseline visit, followed by a 75-g oGTT.

**Fig. 1.**
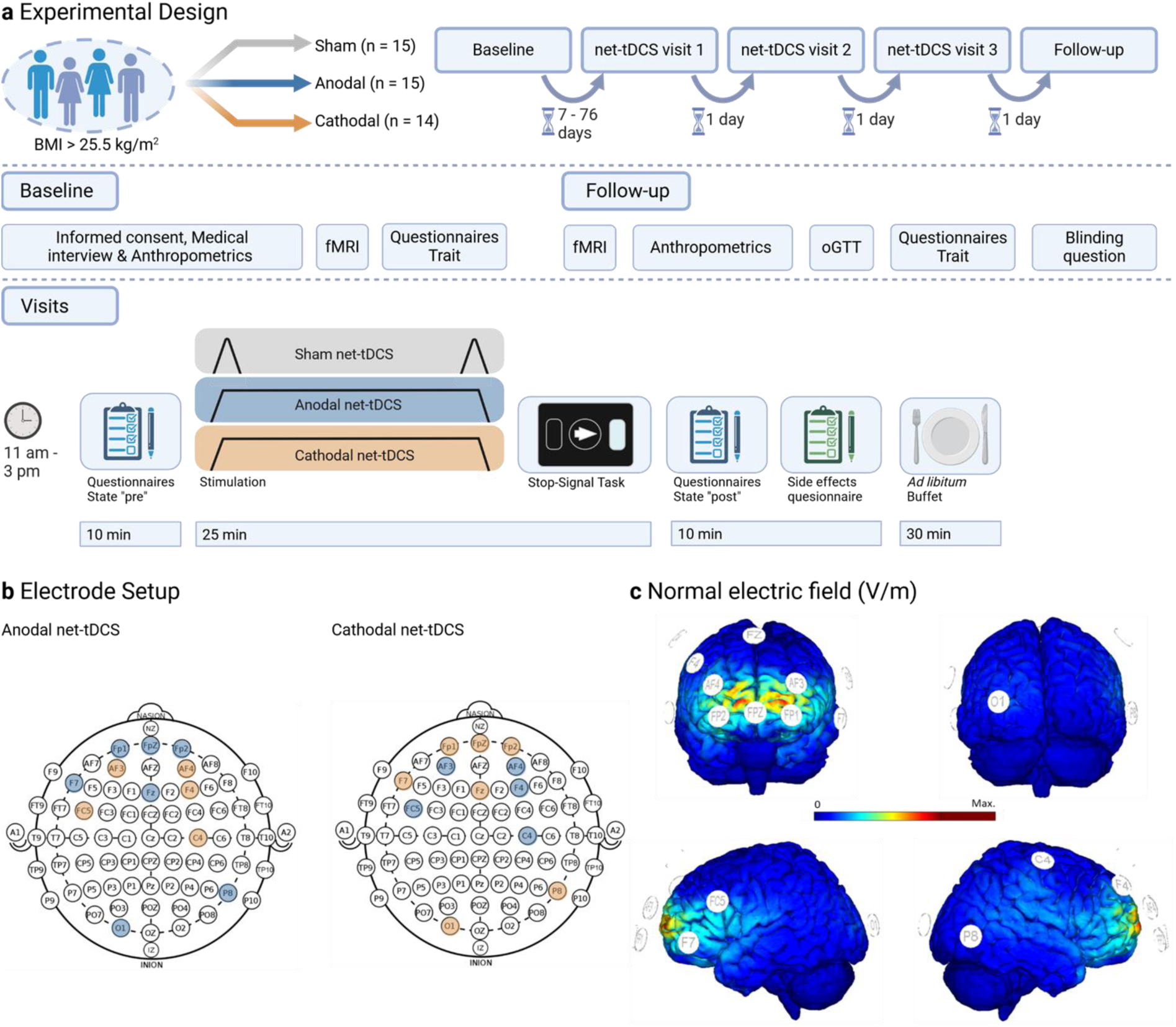
Experimental design, electrode setup and resulting normal electric field. (a) Experimental design. Participants were randomly assigned to either the anodal, cathodal, or sham-net-tDCS group and completed five study days: one baseline assessment, three consecutive visits during which the net-tDCS was administered, and the follow-up. (b) Electrode setup for net-tDCS: The electrode assembly consists of 12 ciruclar Ag/AgCl electrodes. Orange dots show electrodes where the current entered into the cortex; blue dots indicate electrodes where the current dissipated. (c) Normal electric field (V/m) induced by the net-tDCS montage aimed at stimulating the hypothalamus appetite-control network. Figure (c) from [49], originally provided from Neuroelectrics Barcelona S.L.U. Abbreviations: fMRI, functional magnetic resonance imaging; net-tDCS, network-transcranial direct current stimulation; oGTT, oral glucose tolerance test.

### 2.3 Network transcranial direct current stimulation

Stimulation aimed to modulate the hypothalamus appetite control network and stimulation montage was derived as described previously [49]. In brief, electrode position and current strength were determined using the Stimweaver^TM^ algorithm [50], which resulted in a 12-channel optimization. The target network was defined based on the correlation strength of the resting-state FC network of the hypothalamus on the Neurosynth website (https://neurosynth.org/locations/6_2_-10_6/). Stimulation was applied using a network-targeted multichannel tDCS device (Starstim® 32, Neuroelectrics Barcelona S.L.U., Barcelona, Spain) controlled by the corresponding NIC 2.0 software (NIC2 v2.0.11, https://www.neuroelectrics.com/resources/software). Twelve circular Ag/AgCl gelled electrodes were fitted into a neoprene cap (Neoprene Headcap, Neuroelectics Barcelona S.L.U., Barcelona, Spain) at positions defined by the international 10-20 EEG system (see **Suppl. Table 2** for details on electrode placement, current intensities and current density). Prior stimulation, the scalp at the electrode positions was gently rubbed with a cotton swab soaked in alcohol in order to increase current conductivity. Net-tDCS was aimed to activate (anodal) or inhibit (cathodal) the functional connected brain areas of the hypothalamus. Active net-tDCS was delivered for 25 min, including a 15 s ramp-up and 60 s ramp-down at the beginning and end of the session. The maximum total injected current was limited to 4 mA, with each electrode delivering up to 2 mA (in absolute value). A reference electrode for the return current was attached to the participant’s ear. In the sham stimulation, the procedures were identical except that the current was only active during the ramp-up and ramp-down phases using the anodal protocol to simulate the sensation of stimulation. Impedance was kept below 10 kΩ to reduce skin sensation.

#### 2.3.1 Blinding and Tolerability of net-tDCS

Blinding was ensured by using the NIC 2.0 software (Neuroelectrics Barcelona S.L.U., Barcelona, Spain), with both the investigator and participant unaware of the particular protocol being used, which was generically named and activated with a password by a third party in ‘double-blind’ mode. At follow-up, participants were asked what stimulation (active or sham) they believed was delivered during the sessions.

Side effects during net-tDCS were evaluated using a 100 mm VAS directly after the session. Questions included the specification of common possible side effects (itching, tingling, pain, exhaustion and nausea). Moreover, participants were asked on a 100 mm VAS, ‘Overall, how uncomfortable was the stimulation for you?’ (i.e., discomfort). A blank line was included for any other side effects not mentioned as VAS.

### 2.4 Stop-Signal task

For measuring response inhibition during net-tDCS, the SST by CANTAB (Cambridge Neuropsychological Test Automated Battery) was applied simultaneously to the stimulation for 20 minutes.

The Stop Signal Reaction Time (SSRT) was used as the primary outcome parameter. For the task, participants were instructed to press a button on the left or right edge of a tablet computer screen using their corresponding index finger, reacting as swiftly as possible, every time an arrow in the screens’ center pointed to the left or right site. In 25 % of the cases, an additional auditory signal (beep sound) was presented, signalling the participant to withhold the pre-potent response (response inhibition). The experiment includes a 16-item trial block and a total of five test blocks, each block consisting of 64 trials. The task uses a staircase design to dynamically adjust to the participants’ performance, leading to a success rate of 50 % for inhibition. The SSRT is calculated by a stochastic model that takes into account the average reaction time in runs without a stop signal, as well as the time interval between visual and auditory signal in which the participant is still able to successfully inhibit in 50 % of cases [51].

### 2.5 Oral glucose tolerance test

All participants underwent during follow-up an oGTT with a standardized 75 g glucose solution (ACCU-Check® Dextro, Roche, Germany). At six timepoints (pre and 30, 60, 90, 120, and 150 min), blood samples were taken to determine glucose and insulin. Blood glucose was measured via the glucose dehydrogenase method using a HemoCue blood glucose photometer (HemoCueAB, Aengelholm, Sweden). Levels of plasma insulin were determined by using chemiluminesence assays for ADVIA Centaur (Siemens Medical Solutions, Fernwald, Germany).

### 2.6 Ad-libitum food buffet

Ad libitum food consumption was measured using a standardized buffet, which comprised of a vast collection of food items (**Suppl. Table 3** for weight and nutritional information). Participants were instructed to eat as much as they liked within 30 minutes. During this time, they were not allowed to interact with their mobile phone or comparable devices to avoid distractions. Participants were told that they could take left over bakery goods home. Food items were weighted before and after buffet. For calorie intake evaluation, total calorie intake and kilocalories (kcal) intake from marcronutrients were used. Caloric intake by sweet food was separately analyzed, with the analysis being based on the percentage of calories from sweet foods relative to the total kcal intake from each participant. The analysis included the following items: apple filled pastry, chocolate croissant, pastry with with poppy seed filling, cocoa powder for chocolate milk, sugar, apple juice, apple, banana, honey, strawberry jam, hazelnut spread (Nutella), raspberry joghurt, chocolate and vanilla pudding.

### 2.7 Functional magnetic resonance imaging data acquisition

Functional fMRI data of the whole brain were collected at baseline and follow-up using a 3.0 T scanner (Magnetom Prisma, Siemens Healthcare GmbH, Erlangen, Germany) with a 20-channel head coil. Functional resting-state data were obtained by using simultaneous multi– band sequence. The following sequence parameters were used: Acceleration factor = 4; TR = 1.18s; TE = 34ms; FOV = 205 mm2; flip angle 65°; voxel size 2.5 x 2.5 x 2.5 mm^3^; slice thickness 2.5 mm; The images were acquired in an interleaved order. The total scan period was 3 minutes and 56 seconds, with each brain volume consisting of 60 slices and each functional run containing 200 image volumes. Moreover, high-resolution T1 weighted anatomical images (MP2RAGE: 192 slices, matrix: 256 x 240, 1 x 1 x 1 mm^3^) of the brain were acquired.

### 2.8 Resting-state fMRI Data preprocessing and analyses

Analyses of resting state fMRI data were performed using CONN [52] release 22.a [53] and SPM [54] release 12.7771 (see **Suppl. Material**). First-level analysis: Seed-based connectivity maps (SBC) were estimated characterizing the patterns of functional connectivity with the hypothalamus seed and the rest of the brain. The hypothalamus seed was defined as MNI coordinate x = 6, y = 2, z = -10, including a 4 mm sphere according to our previous published research [49]. Functional connectivity strength was represented by Fisher-transformed bivariate correlation coefficients from a weighted general linear model (weighted-GLM), modeling the association between their BOLD signal timeseries. The regions of the hypothalamic FC network were defined based on the hypothalamic resting-state FC pattern (https://neurosynth.org/locations/6_2_-10_6/) thresholded at r < − 0.005 and r > 0.005. The thresholded mask was binarized and SBC with the hypothalamus seed and the mask was computed. The values were extracted from the participant’s first level SBC analysis.

### 2.9 Statistical analysis

Data is presented as mean and standard deviation [SD] unless otherwise stated. Statistical analysis was performed using the program R (R version 4.3.3 (2024-02-29 ucrt), https://cran.r-project.org/bin/windows/base/) for statistical computing and graphics. Statistical significance was set at p ≤ 0.05. The normality of the data distribution was tested using the Shapiro‒Wilk test. When data was not normally distributed, parameters were log-transformed prior analysis or non-parametrical analyses were conducted.

Blinding efficiency as well as differences in side effects between study visits were analyzed using a Friedman-test and Kruskal-Wallis test. For post-hoc comparisons, Dunn-Bonferroni corrections were used.

To compare demographics and scores of psychological questionnaires, an ANOVA or a Kruskal-Wallis test was used. A Fisher’s exact test was examined to compare sex ratio.

To analyze the effect of net-tDCS on eating behavior (state questionnaires and food intake) and response inhibition, we conducted linear mixed effects models (LMER) employing the *lmer* function of the *lme4* package in R. Net-tDCS visits (visit 1, visit 2, visit 3) and net-tDCS group (sham, anodal, cathodal) were entered as fixed effects in all of these models. We used the extracted FC values to evaluate whether changes in resting-state FC of the hypothalamus appetite-control network (follow-up minus baseline) show an interaction with the changes in SSRT. In this exploratory analysis, the changes in hypothalamic FC from baseline to follow-up were considered as fixed effects. Sham and visit 1 were defined as reference category. Participants were included as random factor to account for varying intercepts between participants. To evaluate the effects on peripheral metabolism, we used the *lm* function of the *stats* package in R. Here, condition (sham, anodal, cathodal) were entered as fixed effects in all models and sham was defined as reference category.

To identify the best-fitting model, we employed a stepwise model approach with the Akaike Information Criterion (AIC) as basis for the selection. See **Suppl. Table 4** for the model selection procedures. The general covariates tested across different analyses included age and sex. In more specific analyses, the following covariates were tested: For macronutrient intake, considered covariates were BMI, total kcal intake, and the time period since the last meal (in minutes) prior to net-tDCS. For sweet food intake, the covariates tested were BMI, total kcal intake, and the time period since the last meal (in minutes) prior to net-tDCS. For state questionnaires, baseline values were considered. For total kcal intake, BMI and the time period since the last meal (in minutes) prior to net-tDCS were tested. Residuals of the models were investigated to assess normality and homoscedasticity. To check multicollinearity, we used the *check_collinearity* function in the *performance* package. Results were printed via the *tab_model* function in the *sjPlot* package. Figures were created using the *ggplot2* and *ggeffects* packages.

## 3. Results

### 3.1 Participants

Sixty-six men and women were eligible for the study and forty-four participants were included in the final analysis (36.3 years, BMI 30.6 kg/m^2^, 52.3 % female). The CONSORT flow diagram for the recruitment process is shown in **Suppl. Fig. 1**. Participants’ baseline and anthropometric characteristics are reported in **Table 1**. No significant difference between net-tDCS groups were observed in sex ratio, age, weight, BMI, waist circumference, hip circumference, waist-to-hip ratio (WHR), body-fat content and number of days between baseline and follow-up. In addition, there were no statistical differences in eating behavior trait profiles between stimulation groups (**Suppl. Table 5**).

**Table 1.** Participant baseline and anthropometric characteristics of net-tDCS groups.

|  |  | sham<br>(N=15) | anodal<br>(N=15) | cathodal<br>(N=14) | p-value | Total<br>(N=44) |
| --- | --- | --- | --- | --- | --- | --- |
| **Sex** | Women | 8 (53.3%) | 8 (53.3%) | 7 (50.0%) | 0.979 | 23 (52.3%) |
|  | Men | 7 (46.7%) | 7 (46.7%) | 7 (50.0%) |  | 21 (47.7%) |
| **Age (years)** | Mean (SD) | 36.5 (13.0) | 34.6 (12.0) | 37.8 (15.1) | 0.997 | 36.3 (13.1) |
| **BMI (kg/m<sup>2</sup>)** | Mean (SD) | 30.4 (3.06) | 30.9 (3.86) | 30.6 (2.80) | 0.922 | 30.6 (3.21) |
| **Waist circumference (cm)** | Mean (SD) | 94.0 (8.43) | 95.6 (10.3) | 95.1 (10.3) | 0.902 | 94.9 (9.50) |
| **WHR** | Mean (SD) | 0.880<br>(0.0714) | 0.885<br>(0.0582) | 0.861<br>(0.0866) | 0.651 | 0.876<br>(0.0717) |
| **Body fat content (% , BIA-derived)** | Mean (SD) | 34.4 (10.5) | 34.9 (9.71) | 34.5 (10.2) | 0.999 | 34.6 (9.89) |
| **Number of days between baseline and follow-up visit** | Mean (SD) | 32.2 (21.0) | 24.3 (16.4) | 27.5 (18.0) | 0.611 | 28.0 (18.4) |

Mean values from variables between net-tDCS groups were compared using an ANOVA or a Kruskal-Wallis test. A Fisher's exact test was used to compare sex ratio. Abbreviations: BIA, Bioelectrical Impedance Analysis; BMI, Body Mass Index; WHR, Waist-to-Hip Ratio.

### 3.2 Blinding and tolerability

Participants were not able to distinguish active net-tDCS from sham net-tDCS (χ²(4) = 2.28, p *=* 0.685). **Suppl. Table 6** gives an overview of the net-tDCS groups and the participants’ guess. Active and sham net-tDCS was well tolerated and no participant withdrew from the study due to side effects. There was no significant difference with respect to side effects between net-tDCS groups (see **Suppl. Results** and **Suppl. Table 7** for statistical analysis). We found significant effects of visit; There was a significant difference for the anodal group regarding discomfort (χ²(2) = 7.35, p *=* 0.025) with higher values at visit 2 compared to visit 1 (p_adjust=_ 0.043). Moreover, there was a significant difference in the cathodal group (χ²(2) = 7.00, p *=* 0.030) showing post-hoc higher itching values at visit 1 compared to visit 3 (p_adjust_= 0.024).

### 3.3 Response inhibition

There was a significant main effect of net-tDCS on the SSRT for sham vs. anodal net-tDCS (Estimate = -0.12, 95%, CI [-0.19 – -0.04], p = 0.004) and sham vs. cathodal net-tDCS (Estimate = -0.12, 95%, CI [-0.20 – -0.04], p = 0.005), indicating a greater response inhibition for active net-tDCS. Age was a significant predictor, indicating a higher SSRT with increasing age (Estimate = 0.04, 95%, CI [0.01 – 0.06], p = 0.004). No effects of sex was observed. The raw values of the SSRT for all visits and net-tDCS groups are available in **Suppl. Table 8**.

Since we recently reported that net-tDCS effects on SSRT are associated with hypothalamic FC [49], we investigated in an exploratory analysis whether the change in hypothalamic FC from baseline to follow-up interacts with the net-tDCS induced effect on SSRT. As in the aforementioned model, we found a significant main effect for sham vs. anodal net-tDCS (Estimate = -0.13, 95%, CI [-0.20 – -0.06], p < 0.001) and sham vs. cathodal net-tDCS (Estimate = -0.12, 95%, CI [-0.19 – -0.04], p = 0.002). Moreover, we found a significant interaction for anodal net-tDCS × change in hypothalamic FC (Estimate = -1.27, CI [-2.07 – -0.48], p = 0.002) and cathodal net-tDCS × change in hypothalamic FC (Estimate = -0.67, CI [- 1.31 – -0.03], p = 0.039; **Fig. 2a-c** and **Suppl. Table 9**). This indicates that an increase in hypothalamic FC from baseline to follow-up was related to better response inhibition in the active net-tDCS groups compared to sham.

**Fig. 2.**
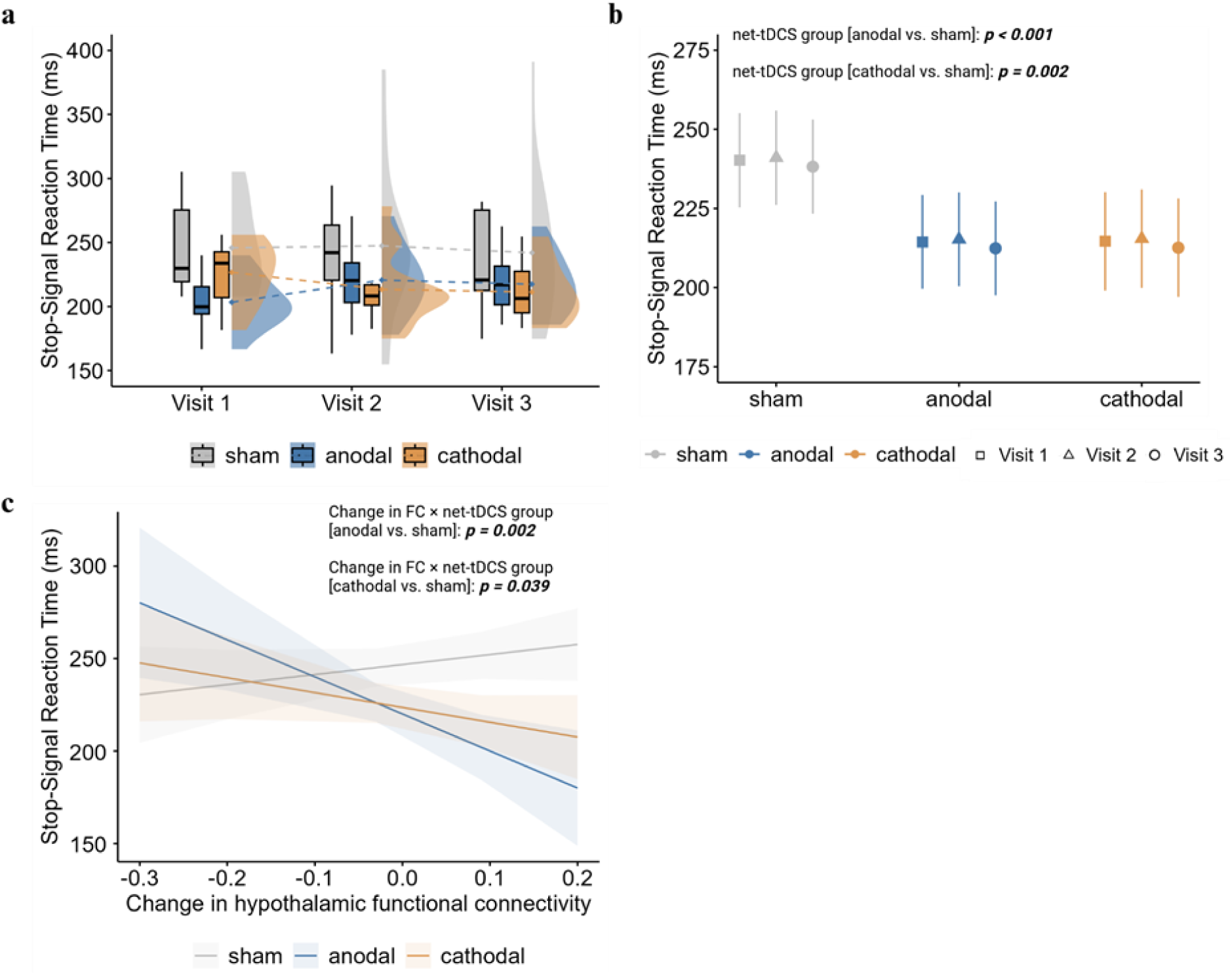
Response inhibition results. **(a)** Raw data of the SSRT for each condition and each visit. Boxplots display median and 1.5 x interquartile range. The half-violin plots show the distribution of the data and the dotted lines display changes of mean values across visits. **(b)** Results indicate a greater response inhibition for active net-tDCS. P values are for main effect of active net-tDCS group compared to sham by two-sided linear mixed model adjusted for age and sex, with change in hypothalamus functional connectivity (FC) as fixed effects. **(c)** Results indicate that a stronger increase in hypothalamic FC from baseline to follow-up is associated with shorter SSRT (better inhibitory control) in the active net-tDCS groups compared to the sham group. P values are for change in hypothalamus FC by net-tDCS group interactions. Shaded areas around the line are depicting the 95% confidence interval. Full results output can be found in **Suppl. Table 9**. N = 44.

### 3.4 Food consumption

No significant main effect of net-tDCS group on total intake (kcal) was observed for sham vs. anodal net-tDCS (Estimate = -108.64, CI = [-519.61 – 302.32], p **=** 0.602) or sham vs. cathodal net-tDCS (Estimate = 72.19, [CI = -346.20 – 490.58], p **=** 0.733); **Suppl. Table 10**). There was no effect of visit. Sex was a significant predictor with male participants showing a higher kcal intake compared to women (Estimate = 752.14, [CI = 412.28 – 1092.00], p < 0.001). Food intake in kcal for all three visits and net-tDCS groups are shown in **Suppl. Table 11 and Fig. 3a-d**.

**Fig. 3.**
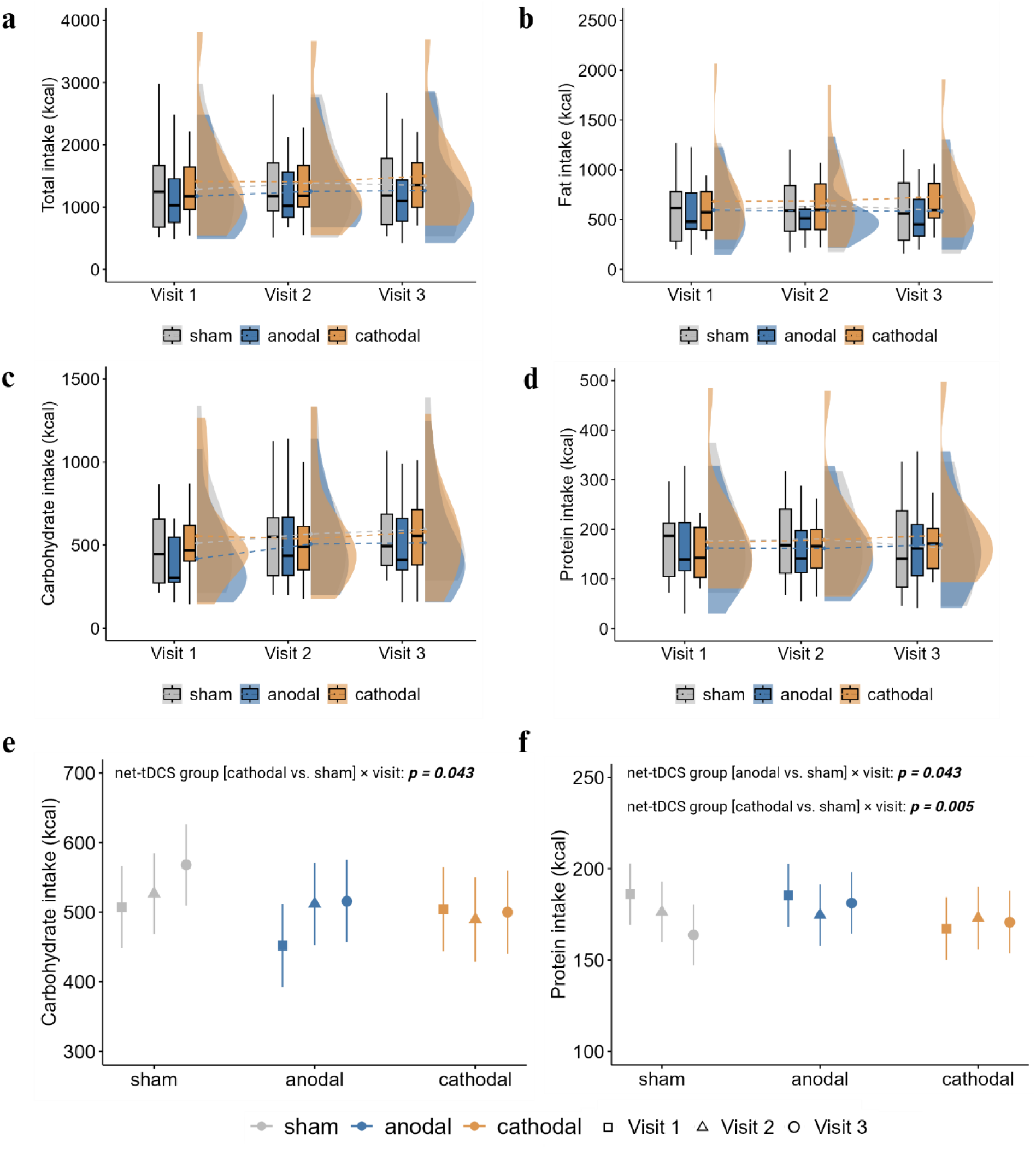
Food consumption results. Raw data of **(a)** total intake, **(b)** fat intake, **(c)** carbohydrate intake and **(d)** protein intake in kcal for each condition and visit. Boxplots display median and 1.5 x interquartile range. The half-violin plots show the distribution of the data and the dotted lines display changes of mean values across visits. **(e-f)** For carbohydrates, participants of the sham group showed a greater consumption during visit 3 compared to visit 1, while the cathodal net-tDCS group showed no increase across visits. For protein consumption, participants of the sham group showed a reduction in protein intake during visit 3 compared to visit 1, while the active net-tDCS groups maintained a consistent protein intake. *P* values are for active net-tDCS group by visit interactions using a two-sided linear mixed model adjusted for sex and total caloric intake. Full results output of all models can be found in **Supp. Table 10, 12**. *N* = 44.

For macronutrient intake, no main effect for sham vs. anodal net-tDCS and sham vs. cathodal net-tDCS was observed (all p > 0.05; **Suppl. Table 12**). For carbohydrate intake, there was a significant interaction effect of cathodal net-tDCS x visit 3 (Estimate = -65.15, [CI = -128.09 – -2.20], p = 0.043, **Fig. 3e**). Hence participants of the sham net-tDCS group consumed more calories during visit 3 compared to visit 1, while the cathodal stimulated group showed no rise in kcal over time. Moreover, for protein intake, there was a significant interaction effect of anodal net-tDCS x visit 3 (Estimate = 18.06, [CI = 0.56 – 35.56], p = 0.043) and cathodal net-tDCS x visit 3 (Estimate = 25.90, [CI = 8.10 – 43.71], p = 0.005, **Fig. 3f**). Hence, participants in the sham group consumed less protein during visit 3 compared to visit 1, while the active tDCS groups showed no reduction in comparison. Anodal tDCS has previously been reported to decrease craving or appetite for sweet foods [33; 35; 36; 38]. Therefore, as an exploratory analysis, we evaluated if sham vs. active net-tDCS was related to lower sweet food intake. Results showed a significant main effect for sham vs. anodal net-tDCS (Estimate = -8.06, [CI = -15.61 – -0.50], p = 0.037; **Fig. 4a-b**) but not for sham vs. cathodal net-tDCS (Estimate = -7.56, [CI = -15.25 – 0.13], p **=** 0.054; **Suppl. Table 13**), indicating a higher sweet food intake for sham vs. anodal but not cathodal net-tDCS. Moreover, total intake (kcal) was a significant predictor (Estimate = 0.01, [CI = 0.00 – 0.01], p = 0.006).

**Fig. 4.**
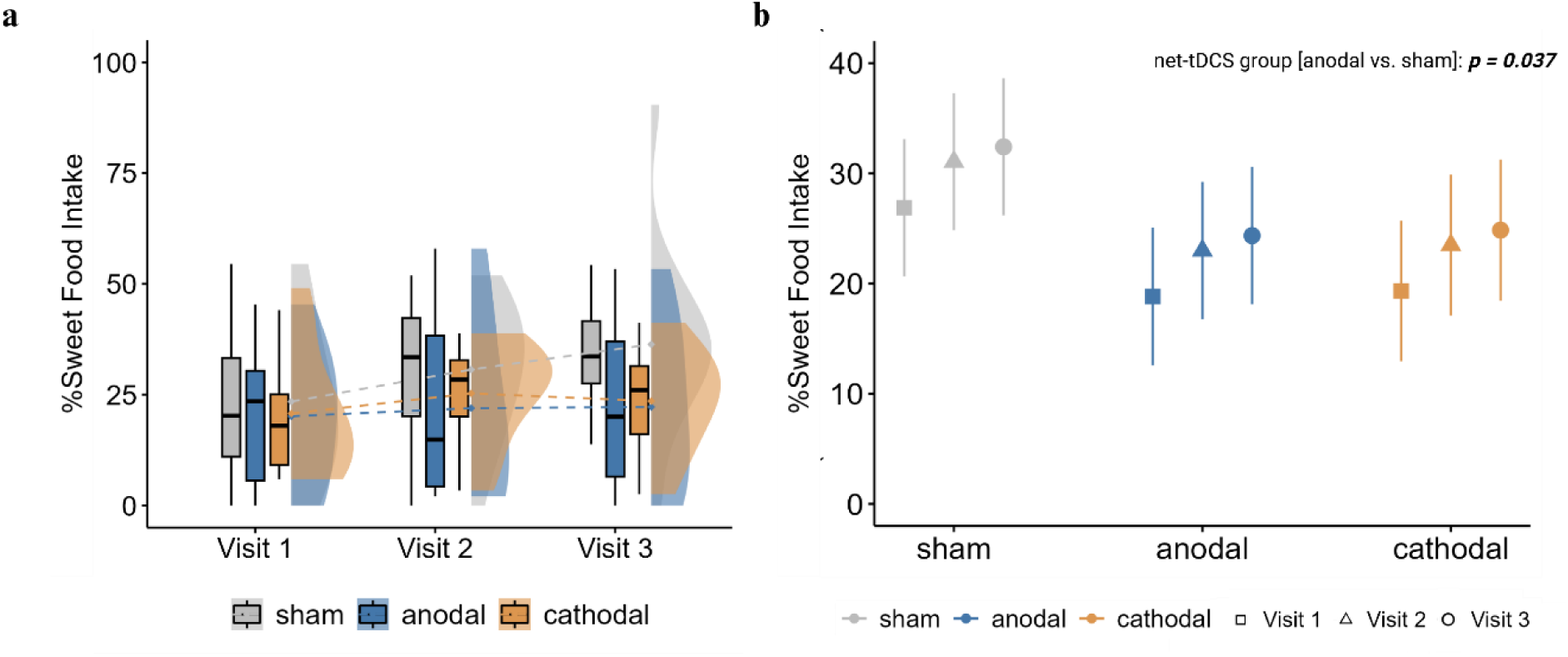
Sweet food intake results. (**a**) Raw data of the percentage of caloric intake by sweet food to total calorie intake for each condition and each visit. Boxplots display median and 1.5 x interquartile range. The half-violin plots show the distribution of the data and the dotted lines display changes of mean values across visits. (**b)** Results show a significantly lower sweet food intake in the anodal net-tDCS group compared to the sham group. *P* value is for main effect of anodal net-tDCS group compared to sham by two-sided linear mixed model adjusted for total caloric intake. Full results output can be found in **Supp. Table 13**. N = 44.

### 3.5 State questionnaires

Baseline ratings for desire to eat, FCQ-S values and PANAS did not differ between groups (see **Suppl. Table 14** for baseline values and analysis). No main effects for sham vs. anodal net-tDCS and sham vs. cathodal net-tDCS for desire to eat ratings, FCQ-S and positive and negative affect of the PANAS questionnaire were found (all p > 0.05; **Suppl. Table 15**).

### 3.6 Glucose metabolism

There was no significant main effect of net-tDCS groups on metabolic parameters (all p > 0.05). The raw metabolic values (insulin sensitivity index (ISI) Matsuda, glycohemoglobin A1c (HbA1c), fasting insulin/glucose, oGTT-derived 2 hour glucose/insulin and triglycerides) divided by net-tDCS groups are shown in **Table 2**. Parameter estimates for the fixed effects, based on the linear models are presented in **Suppl. Table 16**.

**Table 2.** Participant glucose metabolism values at follow-up.

|  |  | **sham**<br>**(N=15)** | **anodal**<br>**(N=15)** | **cathodal**<br>**(N=14)** | **total**<br>**(N=44)** |
| --- | --- | --- | --- | --- | --- |
| **ISI Matsuda,**<br>**oGTT-derived**<br>**(AU)** | Mean (SD) | 12.2 (5.64) | 12.6 (8.80) | 10.5 (3.95) | 11.8 (6.41) |
| **HbA1c**<br>**(mmol/mol)** | Mean (SD) | 34.9 (2.89) | 34.2 (3.75) | 35.3 (4.94) | 34.8 (3.85) |
| **HbA1c (%)** | Mean (SD) | 5.35 (0.261) | 5.29 (0.352) | 5.39 (0.437) | 5.34 (0.349) |
| **Fasting glucose**<br>**(mmol/L)** | Mean (SD) | 4.95 (0.419) | 4.93 (0.363) | 5.16 (0.336) | 5.01 (0.382) |
| **Fasting insulin**<br>**(pmol/L)** | Mean (SD) | 78.7 (28.8) | 83.9 (50.4) | 79.4 (28.6) | 80.7 (36.7) |
| **Glucose, 2 hour**<br>**(mmol/L)** | Mean (SD) | 5.08 (1.19) | 5.16 (1.45) | 5.31 (0.935) | 5.18 (1.19) |
| **Insulin, 2 hour**<br>**(pmol/L)** | Mean (SD) | 338 (389) | 385 (205) | 352 (209) | 359 (277) |
| **Fasting**<br>**Triglycerides**<br>**(mg/dL)** | Mean (SD) | 111 (66.6) | 101 (39.5) | 97.4 (39.9) | 104 (49.7) |

Abbreviations: AU, arbitrary units; HbA1c, glycohemoglobin A1c; ISI, insulin sensitivity index; oGTT, oral glucose tolerance test.

## 4. Discussion

Non-invasive brain stimulation (NIBS) such as tDCS is an emerging tool for improving cognitive control over eating in individuals with overweight or obesity. Unlike earlier studies focusing on single brain areas, net-tDCS targets entire brain networks. Our study examined net-tDCS effects on the hypothalamus appetite-control network. Stimulation was effectively blinded and well tolerated. Our results suggest enhanced inhibitory control and lower caloric intake from sweet food after active net-tDCS compared to sham, with strongest effects after anodal stimulation. Additionally, increased hypothalamic FC predicted better inhibitory control performance in both anodal and cathodal net-tDCS groups.

Overall, our findings imply that especially anodal net-tDCS of the hypothalamus appetite control network can modulate eating behavior and cognitive control. The association between increased hypothalamic FC and enhanced inhibitory control further substantiates the specificity of the stimulation for the intended brain network.

### 4.1 Active net-tDCS related to better inhibitory control

We previously showed in a pilot study an improved response inhibition after anodal net-tDCS compared to sham in a within-subject design in persons with overweight or obesity [49]. Here, we could replicate these effects for anodal net-tDCS, supporting our hypothesis that net-tDCS targeting the hypothalamus appetite-control network can result in better cognitive functions related to inhibitory control. Notably, in contrast to our earlier trial, we also revealed better inhibitory control in the cathodal net-tDCS group compared to sham. In fact, previous research questioned the simple dichotomy of anodal-excitation and cathodal-inhibition effects, which is rarely seen in cognitive studies [55] and cognitive performance was shown to be improved using cathodal tDCS [for overview [56]]. Moreover, Batsikadze et al. [57] demonstrated that low current of 1 mA cathodal tDCS decreased corticospinal excitability while higher current (2 mA) was associated with excitatory effects. The present study stimulated with a total injected current of 4 mA and was aimed to stimulate a brain network, potentially leading to excitatory effects during the cathodal stimulation. Future studies should consider implementing varying levels of injected currents or perform brain imaging measurements to further investigate the effects of different strength of cathodal stimulation.

Resting-state FC was shown to be a predictor for individual variances in cognitive performance and impairments [58; 59]. Thereby, lower resting-state FC in brain regions related to cognition were found in individuals with obesity [9]. Enhancing FC between the dlPFC and ventromedial PFC (vmPFC) was shown to increase food intake-related control mechanisms in participants with overweight or obesity [23], emphasizing the critical role of FC between brain areas in inhibitory control processes. Both the dlPFC and vmPFC are part of the hypothalamic FC network targeted in this study; hence, modulating hypothalamic FC to strengthen control mechanisms might in the long run promote weight loss and support weight maintenance. The current trial showed that higher increase in hypothalamus resting-state FC is associated with better cognitive performance during anodal and cathodal net-tDCS. Evidence from a previous study showed that tDCS can affect FC, which is associated with enhanced response inhibition [60]. In conclusion, our study provides evidence that net-tDCS influences response inhibition with hypothalamus network connectivity playing a crucial role. These findings advance our understanding of the neural mechanisms underlying cognitive enhancement through tDCS.

### 4.2 Anodal net-tDCS does not reduce total caloric intake but associates with lower sweet food intake

In the present study, total kcal intake was not different between anodal, cathodal net-tDCS and sham. This is in contrast to the hypothesized calorie-reducing effects of active stimulations previously reported in overweight and obese participants [35; 39; 61]. For macronutrient intake, however, we found a significant interaction between stimulation group and visit. Active net-tDCS groups maintained a stable intake of protein and carbohydrates over the study visits. The sham group, on the other hand, shifted to an unhealthier eating pattern after repeated exposure to the breakfast buffet, by increasing carbohydrate intake and decreasing protein intake on the third visit. Hence, active net-tDCS may facilitate inhibitory control during exposure to an obesogenic environment. While only few previous studies investigated tDCS effects on macronutrient intake, Jauch-Chara et al. [38] attributed the calorie-reducing effect after anodal tDCS to a reduced carbohydrate intake. Concomitantly, anodal tDCS over the right dlPFC has been previously shown to reduce appetite and cravings for highly palatable foods [33–36; 38], and left dlPFC stimulation decreased actual sweet food intake [39]. On the other hand, tDCS of the right IFG has been linked to increased chocolate consumption [62]. Indeed, variations in stimulation parameters such as stimulation site [63] can contribute to the varied outcomes. In the present study, we observed lower caloric intake by sweet food with anodal, but not cathodal net-tDCS compared to sham. Unlike previous studies focusing on one brain area [43], our design targeted regions functionally connected to the hypothalamus, which is crucial for the regulation of hunger and satiety signals [8]. Stimulation therefore might have followed other mechanisms compared to previous research targeting the dlPFC or IFG solely. In fact, targeting a whole network has been shown to increase excitability twofold compared to bipolar tDCS [47]. Moreover, it has been demonstrated that successful dietary self-control requires the synchronized activation of the dlPFC and vmPFC [23; 24], whereby our target network included both lateral and medial PFC regions. More specifically, the anodal stimulation was aimed to increase excitability of positive functional connections of the hypothalamus as part of the vmPFC, hippocampus and posterior cingulate cortex, while simultaneously decreasing negative functional connections as the striatum and insula cortex. This suggests that stimulating the hypothalamus appetite-control network may more effectively influence hedonic food intake by integrating key brain regions for cognitive control and reward. Further investigation of this specific network is crucial to better understand the underlying processes.

### 4.3 Net-tDCS does not influence subjective rating of food cravings

While previous studies have reported effects of tDCS on food craving [31–36], this was widely questioned in the last years. In the present study, no significant differences were found, which aligns with more recent research. For instance, studies in healthy participants could not show beneficial effects on food craving [64; 65], which was also concluded in a meta-analysis [66]. However, studies in persons with frequent food cravings or persons with binge eating disorders reported reduced food cravings after anodal tDCS [31–36], whereas studies with non-selective samples reported no effect on food craving or desire to eat ratings [67]. Beaumont et al. [68] observed no impact of anodal tDCS over the right dlPFC on craving or appetite in normal and overweight females with mild-to-moderate binge eating, attributing this to the subclinical severity of conditions. In this study, we included individuals with overweight or obesity without eating disorders. Our findings therefore support recent findings suggesting that tDCS does not significantly impact food cravings or desire to eat in a non-clinical population.

### 4.4 Net-tDCS does not affect glucose metabolism 20 hours after last stimulation

In the present study, active net-tDCS did not affect glucose metabolism and peripheral insulin sensitivity, as no effects were detectable at the 20-hour follow-up. During stimulation, previous research has shown improved glucose tolerance. Specifically, anodal tDCS over the primary motor cortex improved systemic glucose tolerance, demonstrated by higher glucose infusion rates (GIRs) derived from a standard hyperinsulinaemic-euglycaemic glucose clamp procedure [40; 42]. Moreover, Kistenmacher et al. [41] found that anodal tDCS lowered blood glucose levels compared to sham, with effects lasting more than 50 minutes post stimulation onset. Given that tDCS is able to increase neuronal excitability [27] and energy levels of the brain [40; 42], it is conceivable that the brain could regulate its own glucose uptake from the periphery as needed. Indeed, a positive correlation has been shown between overall cerebral energy consumption and systemic glucose tolerance [42]. Moreover, the brain and specifically the hypothalamus controls outflows to the periphery that control systemic glucose metabolism [69]. In the present study, the oGTT was conducted more than 20 hours after the third net-tDCS session. Thus, the direct effects of net-tDCS, such as an increased neuronal excitability, are no longer present. Hence, it is not possible to evaluate whether net-tDCS acutely influenced glucose metabolism during stimulation. Further studies are needed to investigate the effects of tDCS on glucose metabolism in a more detailed and time-dependent manner.

### 4.5 Limitations and further directions

In interpreting our findings, several limitations and methodological considerations needs to be acknowledged. First, the present study used a stimulation design, based on fixed electrode positions according to the international 10-20 EEG system. This reflects a standard configuration, individual anatomical differences were not considered. Therefore, it is possible that for some individuals we did not target the optimal stimulation points. Future trials should evaluate the possibility of personalized electrode arrangements, according to their unique brain anatomy and fMRI network dynamics.

Moreover, it is crucial to acknowledge that fMRI measurements and the oGTT were conducted one day after the last visit. Consequently, we cannot decipher direct acute effects of net-tDCS compared to sham on neural activity and functional connectivity as well as metabolism. The excitatory effects of anodal tDCS measured based on changes in motor evoked potential amplitudes haven been shown to be present up to 90 minutes after completion of tDCS [57]. Hence, future studies should implement fMRI recordings directly after stimulation to identify regional and network related changes in response to net-tDCS. Additionally, the time span between baseline and follow-up ranged from 7-76 days which could have influenced outcomes.

Furthermore, hormonal fluctuations, which are known to influence food cravings and brain control on metabolism, as during the menstrual cycle or menopause in women [70; 71], were not taken into account in the present work and should be investigated in future studies.

## 5. Conclusion

In conclusion, these findings demonstrate that anodal and cathodal net-tDCS targeting the hypothalamus appetite-control network is a suitable approach for enhancing inhibitory control. Anodal stimulation shows a greater potential to ameliorate hedonic food intake than cathodal or sham net-tDCS. Furthermore, our results demonstrate the influence of active anodal and cathodal network tDCS on hypothalamic functional connectivity 20 hours after simulation and its potential link to inhibitory control.

## Abbreviations

AIC: Akaike Information Criterion
BMI: Body Mass Index
CANTAB: Cambridge Neuropsychological Test Automated Battery
dlPFC: Dorsolateral Prefrontal Cortex
FC: Functional Connectivity
FCQ-S: Food Craving Questionnaire State
fMRI: Functional Magnetic Resonance Imaging
GIP: Glucose-Dependent Insulinotropic Polypeptide
GIRs: Glucose Infusion Rates
GLM: General Linear Model
GLP1: Glucagon-like Peptide 1
HbA1c: Glycohemoglobin A1c
HD-tDCS: High-definition Transcranial Direct Current Stimulation
IFG: Interior Frontal Gyrus
ISI: Insulin Sensitivity Index
Kcal: Kilocalories
LMER: Linear Mixed Effects Model
mA: Milliampere
MRI: Magnetic Resonance Imaging
Net-tDCS: Network Transcranial Direct Current Stimulation
NIBS: Non-Invasive Brain Stimulation
oGTT: Oral Glucose Tolerance Test
μA: Microampere
PANAS: Positive and Negative Affect Scale
PFC: Prefrontal Cortex
SBC: Seed-Based Connectivity Maps
SD: Standard Deviation
SSRT: Stop-Signal Reaction Time
SST: Stop-Signal Task
T2DM: Type 2 Diabetes Mellitus
tDCS: Transcranial Direct Current Stimulation
VAS: Visual Analogue Scale
WHR: Waist-to-Hip Ratio

## Acknowledgments

We thank all participants for their participation in this project. We thank Ronan Le Gleut, Aaditya Sharma, and Elmar Spiegel from the Core Facility Statistical Consulting at Helmholtz for their statistical consulting. The authors also thank Maike Borutta (University of Tübingen, Tübingen, Germany) for her excellent technical assistance. The study was supported in parts by a grant (01GI0925) from the Federal Ministry of Education and Research (BMBF) to the German Center for Diabetes Research (DZD e.V.). Figures were created with BioRender.com.

## Data availability

The datasets generated and analysed during the current study are not publicly available due to being used in further work which is not yet published, but are available from the corresponding author on reasonable request.

## CRediT authorship contribution statement

TEN: Investigation, Methodology, Visualization, Formal analysis, Data Curation, Writing – original draft, Writing – Review & Editing. RV: Formal Analysis, Methodology, Writing – original draft, Writing – Review & Editing. MB/LT/MG/JT: Investigation, Data Curation. DL: Data Curation. GR/RS: Software, Writing – Review & Editing. MH: Conceptualization, Writing – Review & Editing. ALB: Writing – Review & Editing. CP: Writing – Review & Editing. HP: Conceptualization, Resources, Writing – Review & Editing. SK: Conceptualization, Project administration, Methodology, Validation, Supervision, Funding acquisition, Writing – Review & Editing.

