## Supplementary Information for "Repeated net-tDCS of the hypothalamus appetite-control network reduces inhibitory control and sweet food intake in persons with overweight or obesity"

^1^Institute for Diabetes Research and Metabolic Diseases (IDM) of the Helmholtz Center Munich at the University of Tübingen, Tübingen, Germany; ^2^Department of Internal Medicine, Division of Endocrinology, Diabetology and Nephrology, Eberhard Karls University Tübingen, Tübingen, Germany; ^3^German Center of Diabetes Research (DZD), Tübingen, Germany; ^4^Neuroelectrics Barcelona, Barcelona, Spain; ^5^Institute for Clinical Chemistry and Pathobiochemistry, Department for Diagnostic Laboratory Medicine, Eberhard Karls University Tübingen, Tübingen, Germany; ^6^Division of Endocrinology and Diabetology, Department of Internal Medicine 1, University Hospital Ulm, Ulm, Germany; ^7^Department of Psychiatry and Psychotherapy, Neurophysiology & Interventional Neuropsychiatry, University Hospital Tübingen, Tübingen, Germany; ^8^German Center for Mental Health (DZPG), Partner Site Tübingen, Tübingen, Germany; ^9^Institute of Pharmaceutical Sciences, Department of Pharmacy and Biochemistry; Interfaculty Centre for Pharmacogenomics and Pharma Research at the Eberhard Karls University Tübingen, Tübingen, Germany.

------------

* Corresponding author. Institute for Diabetes Research and Metabolic Diseases (IDM) of the Helmholtz Center Munich at the University of Tübingen. Otfried Müller Str. 47, 72076 Tübingen.

### Suppl. Materials and Methods

#### Suppl. Material and Methods – Reporting checklist

**Suppl. Table 1.** Reporting checklist for tDCS studies based on Buch et al. [1]. Variant appropriate to our study are marked with an “X”.

| **Experimental Design Factors:** | | | |
| --- | --- | --- | --- |
| **Controls used** | □ None | X Sham | □ Active |
| Blinding used | □ None | □ Single | X Double |
| Hypothesis statement | X Yes | □ No |  |
| If Hypothesis-based: |  |  |  |
| Power-analysis statement | □ Yes | X No |  |
| Pre-registration | X Yes | □ No |  |
| Exploratory-based | □ Yes | X No |  |
| Sample-size estimation (i.e. – power analysis) | □ Yes | X No |  |
| **Participant Factors:** | **Reported?** | **Controlled?** |  |
| Number of subjects | X | □ |  |
| Age of subjects | X | □ |  |
| Gender of subjects | X | □ |  |
| Handedness of subjects | □ | □ |  |
| Subjects prescribed medication | □ | X |  |
| Use of CNS active drugs (e.g. anti-convulsants) | □ | X |  |
| Neuropsychological evaluation | □ | X |  |
| Any medical conditions | □ | X |  |
| History of specific repetitive motor activity | □ | □ |  |
| Years of Education completed | □ | X |  |
| **Stimulation Factors:** | **Reported?** | **Controlled?** |  |
| Scalp position of tDCS electrodes | X | □ |  |
| MRI-based localization of tDCS electrodes | □ | □ |  |
| Electrode type (size and geometry) | X | □ |  |
| Current density of applied stimulation | X | □ |  |
| Type of stimulator used (e.g. brand) | X | □ |  |
| Stimulation intensity | X | □ |  |
| Stimulation ramp time | X | □ |  |
| Stimulation duration | X | □ |  |
| Number of Sessions | X | □ |  |
| If Multiple Sessions: |  |  |  |
| Time interval between sessions | X | □ |  |
| Subject attention (level of arousal) during testing | □ | X |  |
| Subject activities during stimulation | X | □ |  |
| tDCS-induced sensations (i.e. – itching, pain, heat, pinching, burning) | X | □ |  |
| **Analysis & Statistics factors:** |  |  |  |
| Effect-size(s) reported | X Yes | □ No |  |
| Raw data uploaded to publicly accessible data repository | □ Yes | X No |  |
| Analyzed data uploaded to publicly accessible data repository | □ Yes | X No |  |
| Full analysis protocol including custom scripts uploaded to publicly accessible data repository | □ Yes | X No |  |

Abbreviations: CNS, central nervous system; MRI, magnetic resonance imaging; net-tDCS; network-transcranial direct current stimulation.

#### Suppl. Material and Methods – Psychometric questionnaires

A number of psychometric questionnaires were used to assess eating behavior traits of participants during the baseline visit. The questionnaires were implemented in the Unipark survey software (Questback AS, Oslo, Norway) and were made available to participants on a tablet computer for electronic completion. General and habitual food cravings were assessed using the 39-item FCQ-T in German [2]. Higher scores suggest more frequent cravings [3]. The German version of the Eating Disorder Examination-Questionnaire (EDE-Q) [4] assesses eating disorder (ED) attitudes and behaviors in four subscales (Restraint, Weight Concern, Shape Concern, and Eating Concern) [5]. In addition, the Three Factor Eating Questionnaire (TFEQ) [6] as a German version [7] was used. The questionnaire examines dimensions of human eating behavior by using three subscales: restraint eating/cognitive restraint of eating, disinhibition and hunger. To measure trait anxiety in general, the stait trait anxiety inventory-trait (STAI-T) [8] was used. Scores range from 20 to 80, with higher values indicating a higher level of anxiety [9]. Moreover, the short version of the Barrat Impulsiveness Scale (BIS-15) [10] in German [11] was used to measure different aspects of trait impulsivity. Finally, to assess out depression, participants performed the German version of the Beck depression inventory II (BDI-II) [12].

#### Suppl. Material and Methods – State questionnaires

###### State food craving

State food craving was assessed using the Food Craving Questionnaire – State (FCQ-S) in the German version [2]. Participants were asked to score in a 15-item version the extent to which they agreed with a statement ‘at the moment’ using a 5-point scale ranging from 1 (*‘strongly disagree’*) to 5 (*‘strongly agree’*). The questionnaire contains five craving subscales namely 1) anticipation of positive reinforcement that may result from eating (pos reinforcement), 2) anticipation of relief from negative feelings as a result of eating (neg reinforcement), 3) desire to eat (desire), 4) lack of control over eating (lack of control) as well as 5) craving as physiological state (hunger). For evaluation, scores were calculated for specific subscales or added up for a total score (ranging from 15 to 75) with higher scores equating to larger momentary craving.

###### Desire to eat

Desire to eat was rated on a 100 mm VAS. Scores range from 0 to 100 whereas higher scores indicate greater desire to eat prevalence.

###### Positive and negative affect schedule

For measuring the positive and negative affect of a person at the moment, the German version of the positive and negative affect schedule (PANAS) [13] was used. The questionnaire contains of two 10-item mood-scales, whereby ten items capture the dimensions of positive and ten of negative affect. Participants were asked to rate every item on a 5-point scale from 1 (‘*not at all*’) to 5 (‘*very much*’). Mean values of the two dimensions were calculated for evaluation.

##### Suppl. Material and Methods – Electrode placement

**Suppl.** **Table 2.** Electrode placement based on the international 10-20 EEG system, applied current and current density for anodal and cathodal net-tDCS. This table was previously published as part of our pilot study [14].

| Electrode placement | Anodal net-tDCS  Applied current [μA] | Anodal net-tDCS  Current density [mA/cm^2^] | Cathodal net-tDCS  Applied current [μA] | Cathodal net-tDCS  Current density [mA/cm^2^] |
| --- | --- | --- | --- | --- |
| P8 | 251 | 0.08 | -251 | -0.08 |
| F4 | -524 | -0.17 | 524 | 0.17 |
| C4 | -353 | -0.11 | 353 | 0.11 |
| FP2 | 345 | 0.11 | -345 | -0.11 |
| FPZ | 2000 | 0.64 | -2000 | -0.64 |
| Fp1 | 342 | 0.11 | -342 | -0.11 |
| AF3 | -1548 | -0.49 | 1548 | 0.49 |
| Fz | 357 | 0.11 | -357 | -0.11 |
| AF4 | -1038 | -0.33 | 1038 | 0.33 |
| O1 | 246 | 0.08 | -246 | -0.08 |
| F7 | 459 | 0.15 | -459 | -0.15 |
| FC5 | -537 | -0.17 | 537 | 0.17 |

Abbreviations: net-tDCS; network-transcranial direct current stimulation; mA, milliampere; μA, microampere.

##### Suppl. Material and Methods – Ad-libitum food buffet

**Suppl. Table 3.** Nutritional information and weight provided for the foods presented in the standardized buffet.

|  | Weight provided (g) | Fat (g) per 100g | Carbohydrates (g)  per 100g | Protein (g)  per 100g | Total  Kcal per 100g |
| --- | --- | --- | --- | --- | --- |
| Bread rolls | ~110 | 1.6 | 46.5 | 7.2 | 236 |
| Wholewheat bread rolls | ~183 | 14.4 | 32.6 | 11.5 | 321 |
| Pretzel | ~144 | 6.6 | 61.4 | 9.4 | 350 |
| Apple filled pastry | ~124 | 15.0 | 47.0 | 4.8 | 342 |
| Chocolate croissant | ~100 | 18.3 | 46.1 | 6.8 | 369 |
| Pastry with poppy seeds | ~128 | 19.1 | 48.3 | 6.1 | 388 |
| Whole milk | ~1000 | 3.5 | 4.7 | 3.5 | 64 |
| Apple juice | ~1000 | 0.0 | 10.9 | 0.0 | 46 |
| Cocoa powder | ~50 | 3.6 | 78.2 | 5.1 | 383 |
| Sugar | ~50 | 0.0 | 100 | 0.0 | 400 |
| Apple | ~300 | 0.0 | 14.0 | 0.0 | 65 |
| Banana | ~400 | 1.0 | 20.0 | 0.0 | 93 |
| Bell pepper | ~200 | 1.0 | 6.0 | 1.0 | 44 |
| Snack tomatoes | ~200 | 1.0 | 3.0 | 0.0 | 20 |
| Cucumber | ~200 | 1.0 | 2.0 | 0.0 | 14 |
| Butter | ~125 | 82.0 | 1.0 | 0.6 | 747 |
| Honey | ~200 | 0.1 | 75.1 | 0.4 | 302 |
| Strawberry Jam | ~50 | 0.2 | 54.1 | 0.5 | 224 |
| Hazelnut spread (Nutella) | ~50 | 30.9 | 57.5 | 6.3 | 539 |
| Cream cheese | ~50 | 21.0 | 4.3 | 5.4 | 226 |
| Cream cheese with herbs | ~45 | 21.0 | 4.4 | 7.2 | 235 |
| Soft cheese | ~105 | 32.0 | 0.5 | 17.0 | 356 |
| Hard cheese | ~112 | 27.5 | < 0.1 | 26 | 352 |
| Salami | ~42 | 28.1 | 1.0 | 24.1 | 355 |
| Ham cold cuts | ~73 | 23.0 | 1.0 | 12.0 | 261 |
| Low-fat natural joghurt (1.5% fat) | ~200 | 1.5 | 6.6 | 5.3 | 64 |
| Raspberry joghurt | ~150 | 2.7 | 14.0 | 3.0 | 92 |
| Chocolate pudding | ~115 | 5.3 | 17.2 | 2.6 | 128 |
| Vanilla pudding | ~115 | 5.4 | 17.3 | 2.2 | 127 |
| Total |  | **366.8** | **774.7** | **168** | **7143** |

Note: Similar products were used when the original products were no longer manufactured. Coffee, hot water for tea and carbonated water were also provided. Calorie and nutritional content was obtained either from the manufacturer or the federal food key (https://blsdb.de/bls?background). Abbreviations: kcal, kilocalories.

##### Suppl. Material and Methods – Resting-state fMRI Data preprocessing and analysis

Pre-processing included slice-timing, realignment, unwarp, normalization into MNI space, segmentation, outlier detection and Gaussian spatial smoothing (FWHM: 6 mm). In addition, functional data were denoised using a standard denoising pipeline [15] including the regression of potential confounding effects characterized by white matter timeseries (5 CompCor noise components), CSF timeseries (5 CompCor noise components), motion parameters and their first order derivatives (12 factors) [16], QC_timeseries regressors (2 components), outlier scans (below 13 factors) [17], session and task effects and their first order derivatives (4 factors), and linear trends (2 factors) within each functional run, followed by bandpass frequency filtering of the BOLD timeseries [18] between 0.008 Hz and 0.1 Hz. CompCor [19; 20] noise components within white matter and CSF were estimated by computing the average BOLD signal as well as the largest principal components orthogonal to the BOLD average, motion parameters, and outlier scans within each subject's eroded segmentation masks. From the number of noise terms included in this denoising strategy, the effective degrees of freedom of the BOLD signal after denoising were estimated to range from 92.3 to 95.5 (average 95.1) across all subjects [21].

#### Suppl. Material and Methods – Statistical analysis

**Suppl. Table 4.** Model selection**.**

| **Model for response inhibition** | | |
| --- | --- | --- |
| **Comparison** | **Model** | **AIC** |
| Null model | ~ 1 + (1 \| id) | -160.22 |
| Fixed effects | ~ net-tDCS group + visit + (1 \| id) | -161.32 |
|  | ~ net-tDCS group + visit + sex + (1 \| id) | -160.18 |
|  | ~ net-tDCS group + visit + age + (1 \| id) | -165.72 |
|  | **~ net-tDCS group + visit + sex + age + (1 \| id)** | **-166.12** |
| Interactions | ~ net-tDCS group*sex + visit + age + (1 \| id) | -163.31 |
| **Model for response inhibition and hypothalamus FC** | | |
| **Comparison** | **Model** | **AIC** |
| Null model | ~ 1 + (1 \| id) | 1288.8 |
| Fixed effects | ~ change in hypothalamus FC*net-tDCS group + visit + (1 \| id) | 1285.6 |
|  | ~ change in hypothalamus FC*net-tDCS group + visit + sex + (1 \| id) | 1287.5 |
|  | **~ change in hypothalamus FC*net-tDCS group + visit + age + (1 \| id)** | **1278.7** |
|  | ~ change in hypothalamus FC*net-tDCS group + visit + sex + age + (1 \| id) | 1279.9 |
| **Model for total kcal** | | |
| **Comparison** | **Model** | **AIC** |
| Null model | ~ 1 + (1 \| id) | 1949.9 |
| Fixed effects | ~ net-tDCS group + visit + (1 \| id) | 1953.9 |
|  | **~ net-tDCS group + visit + sex + (1 \| id)** | **1940** |
|  | ~ net-tDCS group + visit + age + (1 \| id) | 1953.7 |
|  | ~ net-tDCS group + visit + BMI + (1 \| id) | 1954.0 |
|  | ~ net-tDCS group + visit + time period since the last meal (in minutes) + (1 \| id) | 1955.7 |
|  | ~ net-tDCS group + visit + sex + age + (1 \| id) | 1941.1 |
|  | ~ net-tDCS group + visit + sex + BMI + (1 \| id) | 1940.5 |
|  | ~ net-tDCS group + visit + sex + time period since the last meal (in minutes) + (1 \| id) | 1941.9 |
| Interactions | ~ net-tDCS group*visit + sex + (1 \| id) | 1946.2 |
|  | ~ net-tDCS group*sex + visit + (1 \| id) | 1943.8 |
| **Model for carbohydrates** | | |
| **Comparison** | **Model** | **AIC** |
| Null model | ~ 1 + (1 \| id) | 1723.1 |
| Fixed effects | ~ net-tDCS group + visit + (1 \| id) | 1716.7 |
|  | ~ net-tDCS group + visit + sex + (1 \| id) | 1703.2 |
|  | ~ net-tDCS group + visit + age + (1 \| id) | 1717.0 |
|  | ~ net-tDCS group + visit + BMI + (1 \| id) | 1718.2 |
|  | ~ net-tDCS group + visit + time period since the last meal (in minutes) + (1 \| id) | 1718.7 |
|  | ~ net-tDCS group + visit + total intake (kcal) + (1 \| id) | 1565.2 |
|  | ~ net-tDCS group + visit + sex + total intake (kcal) + (1 \| id) | 1565.1 |
|  | ~ net-tDCS group + visit + sex + age + (1 \| id) | 1704.7 |
|  | ~ net-tDCS group + visit + sex + BMI + (1 \| id) | 1705.1 |
|  | ~ net-tDCS group + visit + sex + time period since the last meal (in minutes) + (1 \| id) | 1705.2 |
| Interactions | **~ net-tDCS group*visit + sex + total intake (kcal) + (1 \| id)** | **1564.9** |
|  | ~ net-tDCS group*sex + visit + total intake (kcal) + (1 \| id) | 1568.9 |
| **Model for fat** | | |
| **Comparison** | **Model** | **AIC** |
| Null model | ~ 1 + (1 \| id) | 1804.9 |
| Fixed effects | ~ net-tDCS group + visit + (1 \| id) | 1811.7 |
|  | ~ net-tDCS group + visit + sex + (1 \| id) | 1799.5 |
|  | ~ net-tDCS group + visit + age + (1 \| id) | 1811.4 |
|  | ~ net-tDCS group + visit + BMI + (1 \| id) | 1810.5 |
|  | ~ net-tDCS group + visit + time period since the last meal (in minutes) + (1 \| id) | 1813.6 |
|  | ~ net-tDCS group + visit + total intake (kcal) + (1 \| id) | 1517.8 |
|  | **~ net-tDCS group + visit + sex + total intake (kcal) + (1 \| id)** | **1517.6** |
|  | ~ net-tDCS group + visit + sex + age + (1 \| id) | 1800.6 |
|  | ~ net-tDCS group + visit + sex + BMI + (1 \| id) | 1798.8 |
|  | ~ net-tDCS group + visit + sex + time period since the last meal (in minutes) + (1 \| id) | 1801.5 |
| Interactions | ~ net-tDCS group*visit + sex + total intake (kcal) + (1 \| id) | 1518.9 |
|  | ~ net-tDCS group*sex + visit + total intake (kcal) + (1 \| id) | 1521.5 |
| **Model for protein** | | |
| **Comparison** | **Model** | **AIC** |
| Null model | ~ 1 + (1 \| id) | 1441.9 |
| Fixed effects | ~ net-tDCS group + visit + (1 \| id) | 1449.5 |
|  | ~ net-tDCS group + visit + sex + (1 \| id) | 1437.6 |
|  | ~ net-tDCS group + visit + age + (1 \| id) | 1449.4 |
|  | ~ net-tDCS group + visit + BMI + (1 \| id) | 1448.4 |
|  | ~ net-tDCS group + visit + time period since the last meal (in minutes) + (1 \| id) | 1451.4 |
|  | ~ net-tDCS group + visit + total intake (kcal) + (1 \| id) | 1234.1 |
|  | ~ net-tDCS group + visit + sex + total intake (kcal) + (1 \| id) | 1235.5 |
|  | ~ net-tDCS group + visit + sex + age + (1 \| id) | 1438.7 |
|  | ~ net-tDCS group + visit + sex + BMI + (1 \| id) | 1436.8 |
| Interactions | **~ net-tDCS group*visit + sex + total intake (kcal) + (1 \| id)** | **1232.4** |
|  | ~ net-tDCS group*sex + visit + total intake (kcal) + (1 \| id) | 1238.8 |
| **Model for sweet food intake** | | |
| **Comparison** | **Model** | **AIC** |
| Null model | ~ 1 + (1 \| id) | 1104.6 |
| Fixed effects | ~ net-tDCS group + visit + (1 \| id) | 1103.1 |
|  | ~ net-tDCS group + visit + sex + (1 \| id) | 1100.1 |
|  | ~ net-tDCS group + visit + age + (1 \| id) | 1103.0 |
|  | ~ net-tDCS group + visit + BMI + (1 \| id) | 1103.7 |
|  | ~ net-tDCS group + visit + time period since the last meal (in minutes) + (1 \| id) | 1104.5 |
|  | **~ net-tDCS group + visit + total intake (kcal) + (1 \| id)** | **1098.3** |
|  | ~ net-tDCS group + visit + sex + total intake (kcal) + (1 \| id) | 1099.0 |
|  | ~ net-tDCS group + visit + sex + age + (1 \| id) | 1100.9 |
|  | ~ net-tDCS group + visit + sex + BMI + (1 \| id) | 1099.8 |
|  | ~ net-tDCS group + visit + sex + time period since the last meal (in minutes) + (1 \| id) | 1101.5 |
| Interactions | ~ net-tDCS group*visit + total intake (kcal) + (1 \| id) | 1102.7 |
|  | ~ net-tDCS group*visit + sex + total intake (kcal) + (1 \| id) | 1103.4 |
|  | ~ net-tDCS group*sex + visit + total intake (kcal) + (1 \| id) | 1102.7 |
| **Model for desire to eat ratings** | | |
| **Comparison** | **Model** | **AIC** |
| Null model | ~ 1 + (1 \| id) | 474.13 |
| Fixed effects | ~ net-tDCS group+ visit + (1 \| id) | 472.16 |
|  | ~ net-tDCS group+ visit + sex + (1 \| id) | 473.44 |
|  | ~ net-tDCS group+ visit + age + (1 \| id) | 472.75 |
|  | **~ net-tDCS group+ visit + desire to eat baseline + (1 \| id)** | **467.34** |
|  | ~ net-tDCS group + visit + sex + age + (1 \| id) | 474.33 |
|  | ~ net-tDCS group + visit + sex + desire to eat baseline + (1 \| id) | 468.73 |
| Interactions | ~ net-tDCS group*visit + desire to eat baseline + (1 \| id) | 471.47 |
|  | ~ net-tDCS group*sex + visit + desire to eat baseline + (1 \| id) | 472.07 |
| **Model for FCQ-S ratings** | | |
| **Comparison** | **Model** | **AIC** |
| Null model | ~ 1 + (1 \| id) | 847.34 |
| Fixed effects | ~ net-tDCS group + visit + (1 \| id) | 853.17 |
|  | ~ net-tDCS group + visit + sex + (1 \| id) | 852.71 |
|  | ~ net-tDCS group + visit + age + (1 \| id) | 855.17 |
|  | **~ net-tDCS group + visit + FCQ-S baseline + (1 \| id)** | **836.00** |
|  | ~ net-tDCS group + visit + sex + age + (1 \| id) | 854.60 |
|  | ~ net-tDCS group + visit + sex + FCQ-S baseline + (1 \| id) | 836.91 |
| Interactions | ~ net-tDCS group*visit + FCQ-S baseline + (1 \| id) | 839.31 |
|  | ~ net-tDCS group *sex + visit + FCQ-S baseline + (1 \| id) | 840.22 |
| **Model for PANAS_positive affect_** | | |
| **Comparison** | **Model** | **AIC** |
| Null model | ~ 1 + (1 \| id) | 762.46 |
| Fixed effects | ~ net-tDCS group + visit + (1 \| id) | 764.22 |
|  | ~ net-tDCS group + visit + sex + (1 \| id) | 764.74 |
|  | ~ net-tDCS group + visit + age + (1 \| id) | 763.21 |
|  | **~ net-tDCS group + visit + PANAS_positive affect_ baseline + (1 \| id)** | **738.66** |
|  | ~ net-tDCS group + visit + sex + age + (1 \| id) | 762.54 |
|  | ~ net-tDCS group + visit + sex + PANAS_positive affect_ baseline + (1 \| id) | 740.31 |
| Interactions | ~ net-tDCS group*visit + PANAS_positive affect_ baseline + (1 \| id) | 740.27 |
|  | ~ net-tDCS group*sex + visit + PANAS_positive affect_ baseline + (1 \| id) | 743.75 |
| **Model for PANAS_negative affect_** | | |
| **Comparison** | **Model** | **AIC** |
| Null model | ~ 1 + (1 \| id) | 600.56 |
| Fixed effects | ~ net-tDCS group + visit + (1 \| id) | 604.30 |
|  | ~ net-tDCS group + visit + sex + (1 \| id) | 605.65 |
|  | ~ net-tDCS group + visit + age + (1 \| id) | 605.66 |
|  | ~ net-tDCS group + visit + PANAS_negative affect_ baseline + (1 \| id) | 552.09 |
|  | ~ net-tDCS group + visit + sex + age + (1 \| id) | 607.21 |
|  | ~ net-tDCS group + visit + sex + PANAS_negative affect_ baseline + (1 \| id) | 553.99 |
| Interactions | ~ net-tDCS group*visit + PANAS_negative affect_ baseline + (1 \| id) | 558.09 |
|  | **~ net-tDCS group*sex + visit + PANAS_negative affect_ baseline + (1 \| id)** | **550.96** |
| **Peripheral metabolism** | | |
| **Comparison** | **Model** | **AIC** |
| Null model | ~ 1 + (1 \| id) | 62.0 |
| Fixed effects | **~ net-tDCS group** | **65.7** |
|  | ~ net-tDCS group + sex | 67.6 |
|  | ~ net-tDCS group + age | 67.4 |
|  | ~ net-tDCS group + sex + age | 69.4 |
| Interactions | ~ net-tDCS group*sex + age | 69.9 |

*Note.* Winning models are highlighted in bold. Due to the high multicollinearitiy of the interaction with visit in the model for response inhibition, this interaction was not included in the model. Abbreviations: AIC Akaike Information Criterion.

### Suppl. Results

#### Suppl. Results – Participants

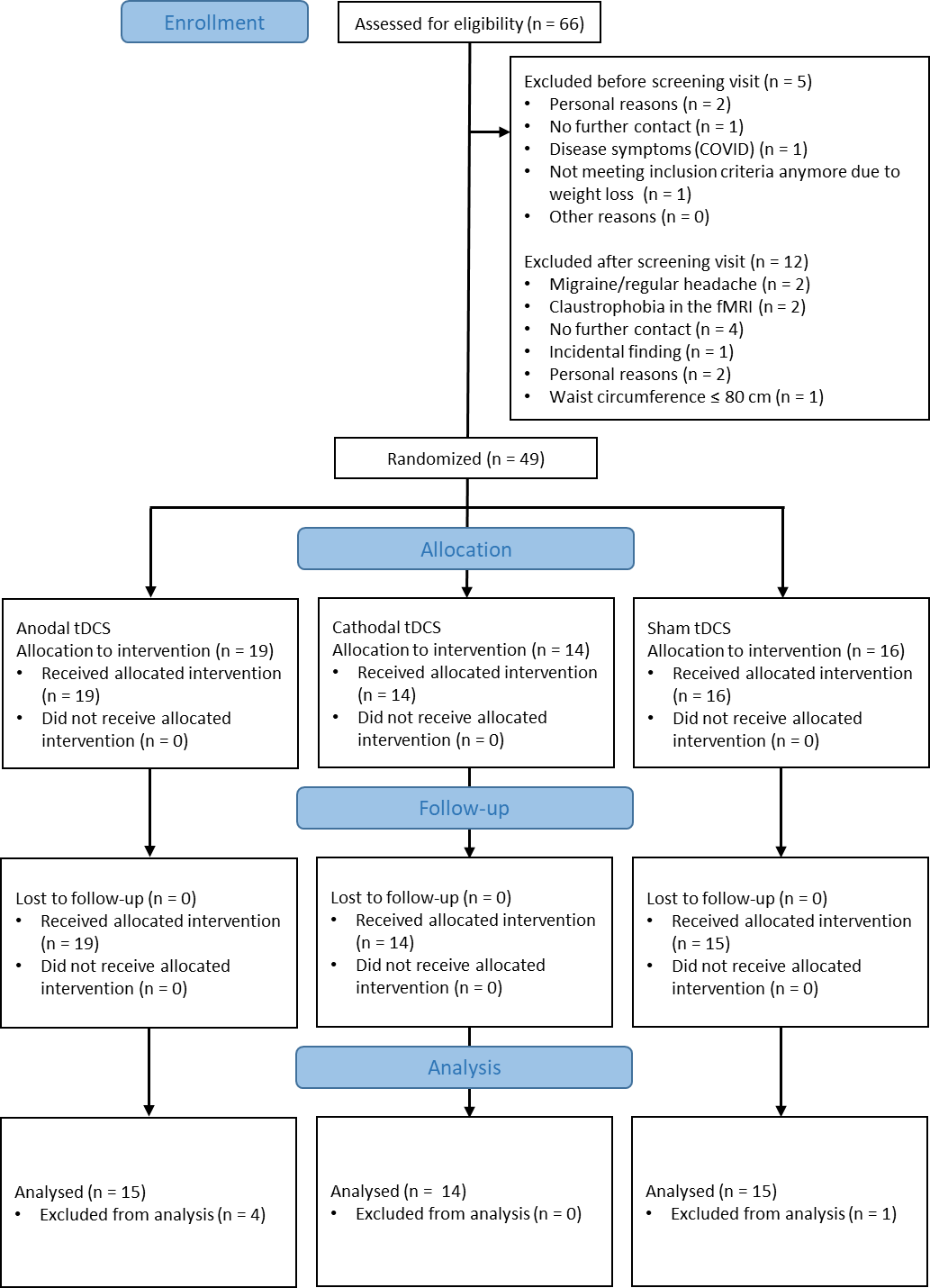

**Suppl. Fig. 1.** CONSORT flow diagram. Initially, 66 participants identified as eligible for the study. Of these, 5 participants did not show up for baseline. Following baseline, 12 individuals withdrew, resulting in a total of 49 participants. All 49 participants completed all 5 study days. For the final analysis, 5 participants were excluded (technical tDCS device issues (n = 2), incidental neurological finding (n = 1), n = 2 more than 5% weight loss after change in lifestyle (n = 1) and death in family (n = 1)), leading to a total of 44 participants for analysis.

#### Suppl. Results – Participants

**Suppl. Table 5.** Mean, standard deviation, median and range for participants eating behaviour trait characteristics of net-tDCS groups and total. Mean values from variables between groups were compared using an ANOVA or a Kruskal-Wallis test.

|  |  |  | sham (N=15) | anodal (N=15) | cathodal (N=14) | p-value | total (N=44) |
| --- | --- | --- | --- | --- | --- | --- | --- |
| FCQ-T | cues | Mean (SD) | 15.4 (4.24) | 13.9 (4.98) | 13.6 (3.15) | 0.485 | 14.3 (4.19) |
|  | control | Mean (SD) | 16.0 (6.27) | 16.7 (8.26) | 16.4 (5.67) | 0.957 | 16.4 (6.69) |
|  | guilt | Mean (SD) | 6.60 (3.64) | 6.80 (3.59) | 6.29 (3.69) | 0.907 | 6.57 (3.56) |
|  | intentions | Mean (SD) | 7.53 (3.38) | 7.93 (4.04) | 8.14 (3.88) | 0.976 | 7.86 (3.70) |
|  | thoughts | Mean (SD) | 13.6 (6.60) | 13.3 (6.93) | 13.8 (6.57) | 0.919 | 13.5 (6.55) |
|  | reinforcement | Mean (SD) | 14.9 (4.67) | 12.7 (5.97) | 11.9 (4.30) | 0.255 | 13.2 (5.10) |
|  | hunger | Mean (SD) | 11.5 (3.91) | 9.67 (5.45) | 9.57 (2.82) | 0.152 | 10.3 (4.22) |
|  | relief | Mean (SD) | 7.27 (3.06) | 7.07 (3.94) | 6.79 (2.91) | 0.877 | 7.05 (3.27) |
|  | emotions | Mean (SD) | 11.9 (4.77) | 10.7 (5.32) | 11.0 (5.67) | 0.831 | 11.2 (5.16) |
|  | total | Mean (SD) | 105 (31.1) | 98.8 (43.0) | 97.6 (27.2) | 0.798 | 100 (33.9) |
| BIS-15 | nonplanning | Mean (SD) | 11.1 (4.06) | 11.4 (3.14) | 10.6 (3.37) | 0.848 | 11.0 (3.48) |
|  | motor | Mean (SD) | 11.1 (3.70) | 11.9 (2.52) | 10.4 (2.41) | 0.396 | 11.2 (2.94) |
|  | attentional | Mean (SD) | 9.13 (3.02) | 9.53 (2.17) | 10.4 (2.34) | 0.226 | 9.68 (2.54) |
|  | total | Mean (SD) | 31.3 (8.43) | 32.9 (6.56) | 31.5 (5.29) | 0.803 | 31.9 (6.79) |
| EDE | restraint | Mean (SD) | 2.07 (1.81) | 1.29 (1.02) | 1.61 (1.42) | 0.667 | 1.66 (1.46) |
|  | eating concern | Mean (SD) | 0.760 (0.741) | 0.867 (1.00) | 0.686 (0.601) | 0.985 | 0.773 (0.789) |
|  | weight concern | Mean (SD) | 2.25 (1.01) | 1.68 (1.40) | 1.97 (0.967) | 0.4 | 1.97 (1.15) |
|  | shape concern | Mean (SD) | 2.53 (1.26) | 2.04 (1.31) | 2.59 (1.28) | 0.453 | 2.38 (1.28) |
|  | mean | Mean (SD) | 1.90 (0.980) | 1.47 (1.07) | 1.72 (0.916) | 0.495 | 1.70 (0.986) |
| BDI | total | Mean (SD) | 6.47 (5.87) | 7.13 (11.4) | 8.71 (6.98) | 0.2 | 7.41 (8.32) |
| TFEQ | restraint | Mean (SD) | 7.27 (5.19) | 5.20 (3.59) | 6.00 (4.30) | 0.462 | 6.16 (4.39) |
|  | disinhibition of control | Mean (SD) | 6.93 (2.52) | 6.20 (3.23) | 6.29 (2.61) | 0.74 | 6.48 (2.77) |
|  | perceived hunger | Mean (SD) | 6.67 (3.24) | 7.07 (3.97) | 5.86 (3.68) | 0.591 | 6.55 (3.59) |
| STAI-T | total | Mean (SD) | 38.5 (8.81) | 37.6 (9.76) | 41.4 (8.62) | 0.502 | 39.1 (9.02) |

Abbreviations: BIS-15, Barratt Impulsiveness Scale-15; BDI, Beck Depression Inventory; BMI, Body Mass Index; EDE, Eating Disorder Examination; FCQ-T, Food Craving Questionnaire - Trait; STAI-T, State-Trait Anxiety Inventory; TFEQ, Three Factor Eating Questionnaire.

#### Suppl. Results – Blinding and tolerability

**Suppl. Table 6.** Blinding Integrity Assessments (percentages within parentheses).

|  |  | Net-tDCS group | | |
| --- | --- | --- | --- | --- |
|  |  | **sham**  **(N=15)** | **anodal**  **(N=15)** | **cathodal**  **(N=14)** |
| Participant’s guess | Sham | 4 (26.7%) | 4 (26.7%) | 4 (28.5%) |
|  | Active | 7 (46.7%) | 9 (60%) | 9 (64.3%) |
|  | Unsure | 4 (26.7%) | 2 (13.3%) | 1 (7.2%) |

Eleven individuals reported further side effects in the blank space of the side effects questionnaire. 1) slight burning sensation under the electrode (1x anodal, 1x cathodal); 2) nausea (1 x cathodal); 3) red spots on the skin at the location of the electrodes (1 x sham); 4) headache (1 x sham); 5) feeling of warmth under the electrode (2 x anodal, 1 x cathodal); 6) fatigue (1 x anodal); 7) more alert and attentive (1 x sham); 8) tingling at the earlobe (1 x anodal).

**Suppl. Table 7.** Mean, standard deviation, median and range for participants’ incidence rates of side effects of net-tDCS separated by group and visit.

|  |  | Visit 1 | | | | | Visit 2 | | | | |
| --- | --- | --- | --- | --- | --- | --- | --- | --- | --- | --- | --- |
|  |  | **sham (N=15)** | **anodal (N=15)** | **cathodal (N=14)** | **χ²** | **p-value** | **sham (N=15)** | **anodal (N=15)** | **cathodal (N=14)** | **χ²** | **p-value** |
| tingling | Mean (SD) | 41.3 (27.8) | 25.3 (25.2) | 45.8 (27.1) |  |  | 29.6 (23.1) | 25.9 (21.0) | 44.4 (26.4) |  |  |
|  | Median [Min, Max] | 43.0 [1.00, 87.0] | 13.0 [2.00, 80.0] | 57.5 [1.00, 82.0] | 4.59 | 0.10 | 21.5 [0, 67.0] | 22.0 [1.00, 63.0] | 49.0 [0, 81.0] | 3.34 | 0.19 |
| itching | Mean (SD) | 17.1 (24.8) | 29.7 (29.2) | 46.6 (33.8) |  |  | 17.2 (18.4) | 23.1 (26.6) | 37.7 (31.6) |  |  |
|  | Median [Min, Max] | 4.00 [0, 87.0] | 26.0 [0, 84.0] | 56.5 [0, 94.0] | 5.69 | 0.06 | 10.5 [0, 56.0] | 16.0 [0, 69.0] | 37.0 [1.00, 82.0] | 4.66 | 0.10 |
| pain | Mean (SD) | 20.4 (26.6) | 13.9 (21.8) | 9.43 (20.4) |  |  | 13.4 (22.4) | 19.1 (30.3) | 10.1 (12.2) |  |  |
|  | Median [Min, Max] | 5.00 [0, 74.0] | 3.00 [0, 76.0] | 2.00 [0, 77.0] | 2.49 | 0.29 | 3.00 [0, 73.0] | 3.50 [0, 100] | 5.00 [0, 39.0] | 0.04 | 0.98 |
| exhaution | Mean (SD) | 24.3 (28.5) | 9.13 (19.0) | 21.1 (23.4) |  |  | 23.4 (26.9) | 16.1 (27.2) | 18.9 (27.6) |  |  |
|  | Median [Min, Max] | 5.00 [0, 83.0] | 1.00 [0, 67.0] | 8.50 [0, 58.0] | 4.64 | 0.10 | 14.0 [0, 74.0] | 1.00 [0, 94.0] | 3.50 [0, 79.0] | 1.84 | 0.40 |
| nausea | Mean (SD) | 2.43 (2.34) | 1.67 (3.03) | 0.818 (1.25) |  |  | 1.54 (2.07) | 3.27 (6.15) | 1.09 (1.30) |  |  |
|  | Median [Min, Max] | 2.50 [0, 7.00] | 0 [0, 10.0] | 0 [0, 4.00] | 2.93 | 0.23 | 0 [0, 6.00] | 1.00 [0, 21.0] | 1.00 [0, 4.00] | 0.53 | 0.77 |
| dis-  comfort^Δ^ | Mean (SD) | 16.5 (16.0) | 17.5 (17.9) | 25.4 (24.2) |  |  | 15.6 (20.1) | 22.5 (22.8) | 21.6 (19.9) |  |  |
|  | Median [Min, Max] | 13.0 [0, 51.0] | 12.0 [0, 61.0] | 16.0 [0, 68.0] | 0.90 | 0.64 | 7.00 [0, 66.0] | 20.0 [0, 62.0] | 12.5 [2.00, 68.0] | 1.81 | 0.40 |

**Continued Suppl. Table 7.**

|  |  | Visit 3 | | | | | Total | | |
| --- | --- | --- | --- | --- | --- | --- | --- | --- | --- |
| tingling | Mean (SD) | **sham (N=15)** | **anodal (N=15)** | **cathodal (N=14)** | **χ²** | **p-value** | **sham (N=15)** | **anodal (N=15)** | **cathodal (N=14)** |
|  | Median [Min, Max] | 37.3 (29.5) | 26.4 (21.7) | 32.5 (26.0) |  |  | 36.2 (26.9) | 25.9 (22.2) | 40.9 (26.5) |
| itching | Mean (SD) | 32.0 [0, 85.0] | 21.0 [1.00, 73.0] | 24.5 [0, 81.0] | 1.07 | 0.59 | 32.0 [0, 87.0] | 20.5 [1.00, 80.0] | 43.0 [0, 82.0] |
|  | Median [Min, Max] | 24.8 (27.9) | 27.7 (28.7) | 27.9 (28.5) |  |  | 19.8 (23.9) | 26.9 (27.7) | 37.4 (31.5) |
| pain | Mean (SD) | 16.0 [0, 78.0] | 23.0 [0, 92.0] | 15.5 [0, 82.0] | 0.18 | 0.91 | 13.0 [0, 87.0] | 22.5 [0, 92.0] | 37.0 [0, 94.0] |
|  | Median [Min, Max] | 23.4 (26.6) | 7.73 (17.4) | 11.0 (14.0) |  |  | 19.2 (25.1) | 13.5 (23.5) | 10.2 (15.6) |
| exhaution | Mean (SD) | 16.0 [0, 76.0] | 1.00 [0, 64.0] | 9.50 [0, 53.0] | 5.30 | 0.07 | 4.50 [0, 76.0] | 2.00 [0, 100] | 4.00 [0, 77.0] |
|  | Median [Min, Max] | 19.9 (25.4) | 9.07 (15.2) | 13.9 (21.8) |  |  | 22.5 (26.4) | 11.3 (20.7) | 18.0 (24.0) |
| nausea | Mean (SD) | 6.00 [0, 70.0] | 1.00 [0, 50.0] | 5.00 [0, 74.0] | 1.36 | 0.51 | 6.50 [0, 83.0] | 1.00 [0, 94.0] | 4.50 [0, 79.0] |
|  | Median [Min, Max] | 2.36 (3.65) | 1.08 (1.88) | 1.75 (3.14) |  |  | 2.12 (2.75) | 1.97 (4.01) | 1.24 (2.10) |
| dis-  comfort^Δ^ | Mean (SD) | 1.00 [0, 12.0] | 0 [0, 5.00] | 1.00 [0, 11.0] | 1.43 | 0.49 | 1.00 [0, 12.0] | 0 [0, 21.0] | 1.00 [0, 11.0] |
|  | Median [Min, Max] | 17.9 (19.3) | 15.9 (19.9) | 22.3 (19.2) |  |  | 16.7 (18.1) | 18.5 (19.9) | 23.1 (20.8) |
|  |  | 10.0 [0, 61.0] | 8.00 [0, 69.0] | 22.0 [1.00, 74.0] | 1.89 | 0.39 | 9.00 [0, 66.0] | 11.5 [0, 69.0] | 16.0 [0, 74.0] |

Discomfort^Δ^, “How uncomfortable was the stimulation for you?”

#### Suppl. Results – Response inhibition

**Suppl. Table 8.** Mean, standard deviation, median and range for participants’ raw Stop-Signal Reaction Time (SSRT) values divided by group and visit.

|  | sham (N=15) | | | anodal (N=15) | | | cathodal (N=14) | | | total (N=44) | | |
| --- | --- | --- | --- | --- | --- | --- | --- | --- | --- | --- | --- | --- |
|  | **Visit 1 (N=15)** | **Visit 2 (N=15)** | **Visit 3 (N=15)** | **Visit 1 (N=15)** | **Visit 2 (N=15)** | **Visit 3 (N=15)** | **Visit 1 (N=14)** | **Visit 2 (N=14)** | **Visit 3 (N=14)** | **Visit 1 (N=44)** | **Visit 2 (N=44)** | **Visit 3 (N=44)** |
| SSRT |  |  |  |  |  |  |  |  |  |  |  |  |
| Mean (SD) | 246 (34.7) | 247 (58.6) | 242 (54.5) | 203 (19.4) | 221 (25.3) | 217 (22.9) | 227 (22.6) | 213 (28.6) | 211 (23.1) | 225 (31.4) | 227 (42.3) | 224 (38.5) |
| Median [Min, Max] | 230 [208, 305] | 242 [155, 385] | 221 [175, 391] | 200 [167, 240] | 220 [178, 271] | 217 [186, 263] | 234 [182, 256] | 208 [175, 278] | 206 [183, 255] | 220 [167, 305] | 220 [155, 385] | 215 [175, 391] |

Abbreviations: SSRT, Stop-Signal Reaction Time.

**Suppl. Table 9.** Results of mixed-effects model for the Stop-Signal Reaction Time (SSRT) derived from the Stop-Signal Task (SST).

|  | **SSRT (log-transf)** | | | **SSRT (log-transf)** | | |
| --- | --- | --- | --- | --- | --- | --- |
| *Predictors* | *Estimates* | *CI* | *p* | *Estimates* | *CI* | *p* |
| (Intercept) | 5.49 | 5.43 – 5.54 | **<0.001** | 5.46 | 5.39 – 5.53 | **<0.001** |
| Change in hypothalamus FC | 0.27 | -0.14 – 0.68 | 0.191 |  |  |  |
| net-tDCS group [anodal] | -0.13 | -0.20 – -0.06 | **<0.001** | -0.12 | -0.19 – -0.04 | **0.004** |
| net-tDCS group [cathodal] | -0.12 | -0.19 – -0.04 | **0.002** | -0.12 | -0.20 – -0.04 | **0.005** |
| sex [men] |  |  |  | 0.05 | -0.01 – 0.12 | 0.119 |
| age | 0.00 | 0.00 – 0.01 | **0.002** | 0.04 | 0.01 – 0.06 | **0.004** |
| visit [Visit 2] | 0.00 | -0.04 – 0.04 | 0.858 | 0.00 | -0.04 – 0.04 | 0.858 |
| visit [Visit 3] | -0.01 | -0.05 – 0.03 | 0.657 | -0.01 | -0.05 – 0.03 | 0.657 |
| Change in hypothalamus FC × net-tDCS group [anodal] | -1.27 | -2.07 – -0.48 | **0.002** |  |  |  |
| Change in hypothalamus FC × net-tDCS group [cathodal] | -0.67 | -1.31 – -0.03 | **0.039** |  |  |  |
| **Random Effects** |  | | |  | | |
| σ2 | 0.01 | | | 0.01 | | |
| τ00 id | 0.01 | | | 0.01 | | |
| ICC | 0.41 | | | 0.47 | | |
| N id | 44 | | | 44 | | |
| Observations | 132 | | | 132 | | |
| Marginal R^2^ / Conditional R^2^ | 0.332 / 0.602 | | | 0.243 / 0.602 | | |

Estimates based on a linear mixed effects model implemented in the “lmer” function of lme4 [22]. Output was obtained using the “tab_model” function from sjPlot [23]. Abbreviations: FC, functional connectivity; SSRT, Stop-Signal Reaction Time.

#### Suppl. Results – Food consumption

**Suppl. Table 10.** Results of mixed-effects models for total food intake (kcal).

|  | **total intake (kcal)** | | |
| --- | --- | --- | --- |
| Predictors | *Estimates* | *CI* | *p* |
| (Intercept) | 943.04 | 607.98 – 1278.11 | **<0.001** |
| net-tDCS group [anodal] | -108.64 | -519.61 – 302.32 | 0.602 |
| net-tDCS group [cathodal] | 72.19 | -346.20 – 490.58 | 0.733 |
| visit [visit 2] | 60.67 | -28.83 – 150.16 | 0.182 |
| visit [visit 3] | 80.62 | -8.88 – 170.11 | 0.077 |
| sex [men] | 752.14 | 412.28 – 1092.00 | **<0.001** |
| **Random Effects** |  | | |
| σ2 | 44980.16 | | |
| τ00 id | 308344.86 | | |
| ICC | 0.87 | | |
| N id | 44 | | |
| Observations | 132 | | |
| Marginal R^2^ / Conditional R^2^ | 0.298 / 0.911 | | |

Estimates based on a linear mixed effects model implemented in the “lmer” function of lme4 [22]. Output was obtained using the “tab_model” function from sjPlot [23].

**Suppl. Table 11.** Mean, standard deviation, median and range for participants’ total kcal intake and kcal intake from protein, carbohydrates and fat divided by group and visit.

|  | Visit 1 | | | | Visit 2 | | | Visit 3 | | | |
| --- | --- | --- | --- | --- | --- | --- | --- | --- | --- | --- | --- |
|  | **sham (N=15)** | **anodal (N=15)** | **cathodal (N=14)** | **sham (N=15)** | | **anodal (N=15)** | **cathodal (N=14)** | | **sham (N=15)** | **anodal (N=15)** | **cathodal (N=14)** |
| total |  |  |  |  | |  |  | |  |  |  |
| Mean (SD) | 1280 (692) | 1180 (615) | 1410 (843) | 1390 (699) | | 1260 (622) | 1400 (813) | | 1350 (759) | 1270 (717) | 1500 (776) |
| Median [Min, Max] | 1250 [516, 2980] | 1030 [489, 2480] | 1170 [546, 3820] | 1180 [508, 2810] | | 1020 [676, 2760] | 1180 [553, 3670] | | 1180 [534, 2840] | 1110 [422, 2860] | 1360 [702, 3690] |
| protein |  |  |  |  | |  |  | |  |  |  |
| Mean (SD) | 176 (89.1) | 162 (80.8) | 173 (104) | 180 (81.4) | | 161 (75.9) | 178 (104) | | 162 (94.3) | 169 (89.1) | 188 (103) |
| Median [Min, Max] | 187 [72.0, 374] | 139 [30.4, 327] | 143 [81.2, 485] | 168 [67.6, 317] | | 141 [55.1, 327] | 166 [63.8, 479] | | 141 [45.7, 337] | 161 [40.7, 357] | 171 [93.5, 498] |
| carbohydrates |  |  |  |  | |  |  | |  |  |  |
| Mean (SD) | 511 (309) | 418 (244) | 554 (315) | 567 (323) | | 506 (272) | 537 (310) | | 595 (318) | 513 (313) | 580 (302) |
| Median [Min, Max] | 446 [213, 1340] | 303 [155, 1080] | 469 [144, 1270] | 549 [200, 1330] | | 436 [199, 1140] | 489 [177, 1340] | | 494 [287, 1390] | 412 [155, 1250] | 556 [160, 1290] |
| fat |  |  |  |  | |  |  | |  |  |  |
| Mean (SD) | 597 (319) | 595 (325) | 684 (452) | 642 (328) | | 588 (317) | 689 (425) | | 593 (367) | 583 (362) | 730 (404) |
| Median [Min, Max] | 616 [202, 1270] | 478 [146, 1230] | 573 [296, 2070] | 589 [173, 1200] | | 512 [219, 1330] | 598 [221, 1850] | | 561 [159, 1210] | 451 [198, 1300] | 595 [317, 1910] |

**Suppl. Table 12.** Results of mixed-effects models for kcal intake from carbohydrates, fat and protein.

|  | **carbohydrate intake (kcal)** | | | **fat intake (kcal)** | | | **protein intake (kcal)** | | |
| --- | --- | --- | --- | --- | --- | --- | --- | --- | --- |
| *Predictors* | *Estimates* | *CI* | *p* | *Estimates* | *CI* | *p* | *Estimates* | *CI* | *p* |
| (Intercept) | 41.86 | -24.75 – 108.47 | 0.216 | -55.13 | -107.08 – -3.18 | **0.038** | 18.12 | -0.87 – 37.11 | 0.061 |
| net-tDCS group [anodal] | -55.02 | -128.03 – 18.00 | 0.138 | 35.11 | -16.14 – 86.35 | 0.178 | -0.56 | -21.38 – 20.25 | 0.957 |
| net-tDCS group [cathodal] | -2.94 | -77.26 – 71.38 | 0.938 | 41.40 | -10.71 – 93.51 | 0.118 | -18.90 | -40.08 – 2.29 | 0.080 |
| visit [visit 2] | 19.50 | -24.40 – 63.40 | 0.381 | -16.99 | -39.83 – 5.85 | 0.143 | -9.73 | -22.15 – 2.69 | 0.123 |
| visit [visit 3] | 60.90 | 17.10 – 104.70 | **0.007** | -32.82 | -55.72 – -9.93 | **0.005** | -22.31 | -34.70 – -9.92 | **0.001** |
| sex [men] | 46.58 | -12.81 – 105.98 | 0.123 | -36.48 | -84.60 – 11.65 | 0.136 | -6.79 | -23.75 – 10.17 | 0.429 |
| total intake (kcal) | 0.35 | 0.31 – 0.39 | **<0.001** | 0.52 | 0.49 – 0.55 | **<0.001** | 0.13 | 0.12 – 0.14 | **<0.001** |
| net-tDCS group [anodal] × visit fac [visit 2] | 40.40 | -21.46 – 102.25 | 0.198 |  |  |  | -1.15 | -18.65 – 16.35 | 0.897 |
| net-tDCS group [cathodal] × visit [visit 2] | -34.03 | -97.11 – 29.05 | 0.288 |  |  |  | 15.53 | -2.31 – 33.37 | 0.087 |
| net-tDCS group [anodal] × visit fac [visit 3] | 2.67 | -59.18 – 64.52 | 0.932 |  |  |  | 18.06 | 0.56 – 35.56 | **0.043** |
| net-tDCS group [cathodal] × visit [visit 3] | -65.15 | -128.09 – -2.20 | **0.043** |  |  |  | 25.90 | 8.10 – 43.71 | **0.005** |
| **Random Effects** | | | |  | | |  | | |

**Continued Suppl. Table 12.**

| σ^2^ | 3658.43 | 2909.04 | 292.77 |
| --- | --- | --- | --- |
| τ_00_ _id_ | 6510.35 | 4036.32 | 533.67 |
| ICC | 0.64 | 0.58 | 0.65 |
| N _id_ | 44 | 44 | 44 |
| Observations | 132 | 132 | 132 |
| Marginal R^2^ / Conditional R^2^ | 0.873 / 0.954 | 0.950 / 0.979 | 0.902 / 0.965 |

Estimates based on a linear mixed effects model implemented in the “lmer” function of lme4 [22]. Output was obtained using the “tab_model” function from sjPlot [23].

**Suppl. Table 13.** Results of mixed-effects models for the percentage of kcal from desserts in relation to the total food intake (kcal).

|  | **desserts and sweets**  **(%corrected for total intake (kcal))** | | |
| --- | --- | --- | --- |
| Predictors | *Estimates* | *CI* | *p* |
| (Intercept) | 18.69 | 10.34 – 27.05 | **<0.001** |
| net-tDCS group [anodal] | -8.06 | -15.61 – -0.50 | **0.037** |
| net-tDCS group [cathodal] | -7.56 | -15.25 – 0.13 | 0.054 |
| visit [visit 2] | 4.17 | -1.37 – 9.71 | 0.139 |
| visit [visit 3] | 5.52 | -0.02 – 11.07 | 0.051 |
| total intake (kcal) | 0.01 | 0.00 – 0.01 | **0.006** |
| **Random Effects** |  | | |
| σ2 | 172.06 | | |
| τ00 id | 51.53 | | |
| ICC | 0.23 | | |
| N id | 44 | | |
| Observations | 132 | | |
| Marginal R^2^ / Conditional R^2^ | 0.151 / 0.347 | | |

Estimates based on a linear mixed effects model implemented in the “lmer” function of lme4 [22]. Output was obtained using the “tab_model” function from sjPlot [23].

#### Suppl. Results – Desire to eat, FCQ-S, PANAS

**Suppl. Table 14.** Mean, standard deviation, median and range for participants’ baseline desire to eat, FCQ-S and PANAS values divided by net-tDCS group and visit.

|  | Visit 1 | | | | Visit 2 | | | | Visit 3 | | | |
| --- | --- | --- | --- | --- | --- | --- | --- | --- | --- | --- | --- | --- |
|  | **sham (N=15)** | **anodal (N=15)** | **cathodal (N=14)** | **p-value** | **sham (N=15)** | **anodal (N=15)** | **cathodal (N=14)** | **p-value** | **sham (N=15)** | **anodal (N=15)** | **cathodal (N=14)** | **p-value** |
| desire to eat (VAS) |  |  |  |  |  |  |  |  |  |  |  |  |
| Mean (SD) | 4.85 (2.92) | 3.87 (2.37) | 5.18 (2.96) |  | 3.73 (2.97) | 3.56 (3.09) | 4.32 (2.91) |  | 3.96 (2.75) | 2.91 (2.78) | 4.92 (2.56) |  |
| Median [Min, Max] | 5.10 [0.100, 9.00] | 3.80 [0.400, 8.60] | 6.40 [0, 8.80] | 0.39 | 4.60 [0, 9.20] | 3.60 [0, 8.80] | 4.25 [0.200, 8.60] | 0.73 | 3.50 [0.400, 9.80] | 2.40 [0, 9.00] | 4.50 [0.100, 9.20] | 0.14 |
| FCQ-S desire |  |  |  |  |  |  |  |  |  |  |  |  |
| Mean (SD) | 8.40 (3.29) | 6.47 (3.46) | 6.79 (2.69) |  | 8.00 (3.46) | 6.27 (3.69) | 7.29 (3.10) |  | 7.79 (3.77) | 5.53 (3.20) | 7.14 (2.85) |  |
| Median [Min, Max] | 9.00 [3.00, 12.0] | 6.00 [3.00, 12.0] | 6.00 [3.00, 12.0] | 0.21 | 7.00 [3.00, 13.0] | 6.00 [3.00, 15.0] | 6.00 [3.00, 12.0] | 0.17 | 7.00 [3.00, 15.0] | 4.00 [3.00, 12.0] | 6.00 [3.00, 12.0] | 0.12 |
| FCQ-S reinforcement |  |  |  |  |  |  |  |  |  |  |  |  |
| Mean (SD) | 7.80 (2.60) | 6.47 (3.04) | 6.14 (2.07) |  | 6.80 (2.54) | 6.40 (2.92) | 5.93 (2.79) |  | 6.71 (2.52) | 5.93 (2.37) | 6.29 (3.02) |  |
| Median [Min, Max] | 8.00 [3.00, 11.0] | 7.00 [3.00, 12.0] | 7.00 [3.00, 10.0] | 0.20 | 7.00 [3.00, 11.0] | 6.00 [3.00, 11.0] | 6.00 [3.00, 11.0] | 0.73 | 6.00 [3.00, 11.0] | 6.00 [3.00, 10.0] | 6.50 [3.00, 12.0] | 0.75 |
| FCQ-S relief |  |  |  |  |  |  |  |  |  |  |  |  |
| Mean (SD) | 7.00 (2.33) | 5.93 (3.20) | 6.21 (2.08) |  | 6.33 (2.41) | 6.27 (2.81) | 5.43 (1.87) |  | 6.00 (2.25) | 5.87 (2.75) | 5.93 (2.27) |  |
| Median [Min, Max] | 7.00 [3.00, 10.0] | 4.00 [3.00, 11.0] | 6.00 [3.00, 10.0] | 0.43 | 6.00 [3.00, 12.0] | 6.00 [3.00, 12.0] | 5.50 [3.00, 9.00] | 0.63 | 6.00 [3.00, 10.0] | 6.00 [3.00, 11.0] | 6.00 [3.00, 9.00] | 0.95 |
| FCQ-S control |  |  |  |  |  |  |  |  |  |  |  |  |
| Mean (SD) | 6.80 (2.73) | 5.40 (2.56) | 5.93 (1.54) |  | 6.13 (2.70) | 5.13 (2.56) | 5.29 (1.68) |  | 5.14 (2.07) | 5.13 (2.42) | 5.00 (1.71) |  |
| Median [Min, Max] | 7.00 [3.00, 12.0] | 5.00 [3.00, 12.0] | 5.50 [4.00, 10.0] | 0.17 | 6.00 [3.00, 11.0] | 4.00 [3.00, 11.0] | 5.00 [3.00, 10.0] | 0.48 | 5.00 [3.00, 9.00] | 4.00 [3.00, 11.0] | 5.00 [3.00, 9.00] | 0.96 |
| FCQ-S total |  |  |  |  |  |  |  |  |  |  |  |  |

**Continued Suppl. Table 14.**

| Mean (SD) | 40.1 (10.2) | 31.5 (13.5) | 33.9 (7.13) |  | 36.2 (11.6) | 31.6 (11.5) | 31.9 (8.41) |  | 33.6 (10.0) | 29.3 (10.8) | 33.1 (9.83) |  |
| --- | --- | --- | --- | --- | --- | --- | --- | --- | --- | --- | --- | --- |
| Median [Min, Max] | 42.0 [15.0, 51.0] | 32.0 [15.0, 56.0] | 32.5 [22.0, 47.0] | 0.07 | 35.0 [16.0, 55.0] | 31.0 [15.0, 54.0] | 30.5 [22.0, 49.0] | 0.41 | 31.5 [17.0, 48.0] | 29.0 [15.0, 47.0] | 35.0 [19.0, 51.0] | 0.44 |
| PANAS_positive affect_ |  |  |  |  |  |  |  |  |  |  |  |  |
| Mean (SD) | 28.8 (4.89) | 27.6 (5.67) | 30.4 (4.55) |  | 29.5 (5.94) | 24.1 (6.23) | 27.4 (6.74) |  | 25.2 (7.20) | 24.5 (6.59) | 27.4 (6.20) |  |
| Median [Min, Max] | 29.0 [22.0, 37.0] | 27.0 [18.0, 39.0] | 29.5 [24.0, 39.0] | 0.35 | 29.0 [21.0, 45.0] | 24.0 [14.0, 36.0] | 26.0 [17.0, 39.0] | 0.07 | 24.0 [13.0, 45.0] | 23.0 [15.0, 35.0] | 26.5 [18.0, 40.0] | 0.38 |
| PANAS_negative affect_ |  |  |  |  |  |  |  |  |  |  |  |  |
| Mean (SD) | 13.3 (5.00) | 13.2 (3.63) | 13.9 (3.59) |  | 12.3 (3.65) | 11.9 (2.19) | 13.3 (3.29) |  | 11.9 (3.18) | 12.2 (3.00) | 11.9 (1.44) |  |
| Median [Min, Max] | 12.0 [10.0, 26.0] | 12.0 [10.0, 22.0] | 13.5 [10.0, 22.0] | 0.62 | 10.0 [10.0, 20.0] | 11.0 [10.0, 18.0] | 12.0 [10.0, 22.0] | 0.22 | 10.0 [10.0, 21.0] | 11.0 [10.0, 21.0] | 12.0 [10.0, 15.0] | 0.37 |

Abbreviations: FCQ-S, Food Craving Questionnaire – State; PANAS, Positive and negative affect scale; VAS, Visual Analogue Scale.

**Suppl. Table 15.** Results of mixed-effects models for Δ desire to eat, Δ FCQ-S total, Δ positive affect and Δ negative affect of the PANAS questionnaire. Delta (Δ) stands for the change in the respective variable from pre net-tDCS to post net-tDCS measurements.

|  | **Δ desire to eat** | | | **Δ FCQ-S total** | | | **Δ PANAS_positive affect_** | | | **Δ PANAS_negative affect_** | | |
| --- | --- | --- | --- | --- | --- | --- | --- | --- | --- | --- | --- | --- |
| *Predictors* | *Estimates* | *CI* | *p* | *Estimates* | *CI* | *p* | *Estimates* | *CI* | *p* | *Estimates* | *CI* | *p* |
| (Intercept) | 0.98 | 0.25 – 1.71 | **0.009** | 12.12 | 6.65 – 17.58 | **<0.001** | 8.81 | 4.28 – 13.34 | **<0.001** | 5.36 | 3.55 – 7.16 | **<0.001** |
| net-tDCS group [anodal] | -0.05 | -0.73 – 0.63 | 0.887 | -2.27 | -5.99 – 1.46 | 0.231 | 1.69 | -1.09 – 4.48 | 0.231 | 1.32 | -0.23 – 2.87 | 0.094 |
| net-tDCS group [cathodal] | -0.47 | -1.15 – 0.22 | 0.179 | -1.82 | -5.56 – 1.91 | 0.336 | 1.49 | -1.32 – 4.29 | 0.298 | -0.21 | -1.82 – 1.40 | 0.794 |
| visit [Visit 2] | 0.31 | -0.23 – 0.85 | 0.260 | 0.08 | -1.86 – 2.02 | 0.935 | -0.67 | -1.96 – 0.62 | 0.304 | -0.54 | -1.19 – 0.12 | 0.106 |
| visit [Visit 3] | 0.64 | 0.09 – 1.19 | **0.024** | 0.68 | -1.31 – 2.68 | 0.501 | -0.33 | -1.67 – 1.02 | 0.632 | -0.03 | -0.70 – 0.65 | 0.932 |

**Continued Suppl. Table 15.**

| baseline value | -0.13 | -0.23 – -0.03 | **0.008** | -0.29 | -0.41 – -0.17 | **<0.001** | -0.43 | -0.56 – -0.29 | **<0.001** | -0.48 | -0.59 – -0.37 | **<0.001** |
| --- | --- | --- | --- | --- | --- | --- | --- | --- | --- | --- | --- | --- |
| sex [men] |  |  |  |  |  |  |  |  |  | 0.11 | -1.51 – 1.73 | 0.893 |
| net-tDCS group [anodal] × sex [men] |  |  |  |  |  |  |  |  |  | -1.54 | -3.81 – 0.73 | 0.182 |
| net-tDCS group [cathodal] × sex [men] |  |  |  |  |  |  |  |  |  | 1.71 | -0.59 – 4.02 | 0.144 |
| **Random Effects** | | | |  | | |  | | |  | | |
| σ^2^ | 1.62 | | | 20.84 | | | 8.93 | | | 2.34 | | |
| τ_00_ _id_ | 0.32 | | | 18.34 | | | 11.56 | | | 1.65 | | |
| ICC | 0.16 | | | 0.47 | | | 0.56 | | | 0.41 | | |
| N _id_ | 44 | | | 44 | | | 44 | | | 44 | | |
| Observations | 130 | | | 130 | | | 130 | | | 130 | | |
| Marginal R^2^ / Conditional R^2^ | 0.131 / 0.274 | | | 0.196 / 0.572 | | | 0.278 / 0.685 | | | 0.443 / 0.673 | | |

Estimates based on a linear mixed effects model implemented in the “lmer” function of lme4 [22]. Output was obtained using the “tab_model” function from sjPlot [23].

#### Suppl. Results – Peripheral metabolism

**Suppl. Table 16.** Results of mixed-effects models for ISI Matsuda, NEFA-ISI, HbA1c and fasting glucose.

|  | **ISI Matsuda, OGTT-derived (AU)** | | | **NEFA-ISI, OGTT-derived (AU)** | | | **HbA1c (%)** | | | **Fasting glucose (mmol/L)** | | |
| --- | --- | --- | --- | --- | --- | --- | --- | --- | --- | --- | --- | --- |
| *Predictors* | *Estimates* | *CI* | *p* | *Estimates* | *CI* | *p* | *Estimates* | *CI* | *p* | *Estimates* | *CI* | *p* |
| (Intercept) | 2.39 | 2.14 – 2.64 | <0.001 | 1.22 | 1.03 – 1.41 | <0.001 | 1.68 | 1.64 – 1.71 | **<0.001** | 4.95 | 4.75 – 5.14 | **<0.001** |
| net-tDCS group [anodal] | -0.02 | -0.38 – 0.33 | 0.905 | 0.01 | -0.26 – 0.28 | 0.922 | -0.05 | -0.32 – 0.21 | 0.686 | -0.02 | -0.30 – 0.26 | 0.893 |
| net-tDCS group [cathodal] | -0.10 | -0.47 – 0.26 | 0.566 | -0.06 | -0.34 – 0.21 | 0.635 | 0.04 | -0.23 – 0.31 | 0.769 | 0.21 | -0.07 – 0.50 | 0.132 |
| **Random Effects** | | | |  | | |  | | |  | | |
| Observations | 44 | | | 44 | | | 43 | | | 44 | | |
| Marginal R^2^ / Conditional R^2^ | 0.009 / -0.039 | | | 0.009 / -0.039 | | | 0.012 / -0.037 | | | 0.077 / 0.032 | | |

Estimates based on a linear effects model implemented in the “lm” function of lme4 [22]. Output was obtained using the “tab_model” function from sjPlot [23]. Abbreviations: AU, arbitrary units; CI, Confidence interval; df = degrees of freedom; HbA1c, glycohemoglobin A1c; HDL, high-density lipoprotein; ISI, insulin sensitivity index; LDL, low-density lipoprotein; NEFA, nonesterified fatty acid; OGTT, oral glucose tolerance test.
